# Deep Learning Trained Without Target-Domain Labels Improves Risk Stratification Over Expert Coronary Calcium Reading on Routine Attenuation Correction CT

**DOI:** 10.64898/2026.07.02.26356002

**Authors:** Aakash Shanbhag, Robert JH Miller, Aditya Killekar, Anna M Marcinkiewicz, Jianhang Zhou, Mark Lemley, Assiata Kamagate, Serge D. Van Kriekinge, Paul B. Kavanagh, Attila Feher, Edward J Miller, Joanna X. Liang, Daniel S. Berman, Damini Dey, Richard M Leahy, Piotr J. Slomka

**Affiliations:** Artificial Intelligence in Medicine Research Center, Departments of Biomedical Sciences, Medicine, and Cardiology, Cedars-Sinai Medical Center, Los Angeles, CA, USA; Signal and Image Processing Institute, Ming Hsieh Department of Electrical and Computer Engineering, University of Southern California, Los Angeles, California, United States; Department of Cardiac Sciences, University of Calgary, Calgary AB, Canada; Center of Radiological Diagnostics, National Medical Institute of the Ministry of the Interior and Administration, Warsaw, Poland; Section of Cardiovascular Medicine, Department of Internal Medicine, Yale University School of Medicine, New Haven, CT, USA

## Abstract

Coronary artery calcium (CAC) is an established measure of coronary atherosclerosis on computed tomography (CT). Many SPECT and most PET myocardial perfusion studies already include a low-dose CT scan from which coronary calcium could be reported, yet it almost never is. CAC may not be reported due to image noise, cardiac motion, or the absence of a reliable reference standard. Deep learning (DL) models trained on these low-dose CT scans inherit the same uncertainty, because their reference labels are drawn on the degraded images. We bypassed this limitation by training a DL CAC model entirely outside the target domain: expert labels from high-quality gated CT were inherited by synthetic ungated images derived from those same scans, so that no CTAC scan was annotated, and no real CTAC image was used, at any point in training. Models were trained using one center and externally tested in two other centers. Because each synthetic image is derived from the gated scan itself, expert gated segmentations remain valid voxel-for-voxel without registration. Using these pairs, we developed a cross-domain knowledge adaptation model (knowledge adaptation, KAD-CAC). The DL models were externally tested on 5969 patients with a median age of 64 (IQR 56 – 73), of whom 50.2% were male. The KAD-CAC model had higher Cohen’s kappa K (0.86, 95% CI 0.84 – 0.88) compared to a convolutional LSTM model (K 0.78, 95% CI 0.76 – 0.80, p<0.01), or models trained with only gated images (K 0.81, 95% CI 0.80 – 0.82, p<0.01). Net reclassification improvement for death or myocardial infarction over a clinical and perfusion baseline was greatest for the KAD-CAC model and exceeded that of expert CAC reading on the same CTAC scans (difference 0.089, 95% CI 0.051 to 0.125, p<0.001). A model trained without any labels from CTAC scans therefore stratified risk of death or MI better than expert reading of the same scans. Patients classified as CAC 0 by KAD-CAC had lower adjusted risk of death or MI than patients classified as CAC 0 by experts (HR 0.50, 95% CI 0.25 – 0.98). We use paired synthetic ungated scans to transfer expert gated CAC annotations into the ungated domain, resulting in better vessel-level CAC scoring and improved risk reclassification compared with expert reading of the same scans. Automated CAC scoring could be added to every myocardial perfusion imaging report without additional imaging or radiation. The approach provides a general route to expert-quality supervision for any acquisition in which the native reference standard is unreliable.

## Introduction

Coronary artery disease (CAD) is the leading cause of morbidity and mortality worldwide (1). Accurate risk stratification is critical for guiding medical therapy for CAD. Coronary artery calcification (CAC) is an established marker of coronary atherosclerosis (2). The presence of CAC is associated with an increased risk of future cardiovascular events in both symptomatic and asymptomatic patients (3, 4). In addition, the vascular distribution of CAC has been shown to have important prognostic implications beyond the global CAC score (5, 6). CAC was originally described as a measure on dedicated, ECG-gated, computed tomography (CT) scans. However, there is now extensive literature demonstrating that CAC can also be evaluated on low dose lung cancer screening (7), and ungated CT attenuation correction (AC) imaging (8–10). CTAC scans are acquired with every SPECT and PET myocardial perfusion study, yet CAC on these scans is seldom reported in routine practice, so prognostic information already present in the imaging is discarded.

Deep learning (DL) has emerged as an efficient method to automate quantification of CAC on CT. Previous studies have shown the ability of a variety of DL models to identify CAC (11). It was previously demonstrated that a convolutional Long Short-Term Memory model (cLSTM) can accurately quantify CAC (8, 9). However, the low-dose CTAC scans acquired with SPECT and PET have relatively high image noise. A typical radiation dose associated with a CTAC scan is 0.2 to 0.3 mSv, while a typical radiation dose for a dedicated CAC scan is 1 to 3 mSv (12). This is further compounded by the lack of cardiac gating which results in artifacts related to coronary motion. As a result, it can be difficult to accurately identify small lesions and correctly assign vessel-specific CAC even for expert clinicians. Although prior DL approaches have demonstrated the feasibility of CAC quantification on CTAC for total CAC, they have limited accuracy at the lesion level. A central reason is that these models are trained against reference annotations drawn on the same degraded images they are meant to interpret, so the supervision signal itself inherits the reader uncertainty it is intended to overcome. Errors therefore concentrate precisely where expert ungated annotation is least reproducible: small lesions, and vessels most affected by coronary motion

The aim of this study was to develop and externally test a training strategy that supplies expert-quality supervision for CAC segmentation on ultra-low dose CTAC scans. Rather than annotating degraded images, we generate synthetic low-dose ungated images directly from gated CAC scans so that model training can utilize expert segmentations from dedicated gated CAC scans as ground truth while also learning to discern image noise and motion artifacts from low-dose ungated CT. We evaluated agreement with expert CAC segmentations as well as risk stratification in an external population.

## Results

### Study population

The DL model was externally tested on 5969 patients with a median age of 64 (IQR 56 – 73), of whom 2995 (50.2%) were male. Characteristics of the training and external testing sets are shown in **Table 1**. A summary of CAC scores using the various models is shown in **Supplementary Table 1.** Training and internal validation summary of CAC scores in outlined in **Supplementary Table 2.**

**Table 1.** Population characteristics in the training and external testing datasets. Continuous variables shown as median (interquartile range)

|  | **Overall**<br>**N=8335** | **Training and**<br>**Validation**<br>**N=2366** | **External testing**<br>**N=5969** | **P-value** |
| --- | --- | --- | --- | --- |
| Age | 66 (57, 74) | 70 (62, 78) | 64 (56, 73) | <0.001 |
| Male (%) | 4160 (49.9%) | 1165 (49.2%) | 2995 (50.2%) | 0.44 |
| Body mass index<br>(kg/m<sup>2</sup>) | 28.7 (25, 33.3) | 27.6 (24.2, 32) | 29.2 (25.4, 33.8) | <0.001 |
| Hypertension | 5285 (63.4%) | 1744 (73.7%) | 3541 (59.3%) | <0.001 |
| Diabetes mellitus | 2160 (25.9%) | 719 (30.4%) | 1441 (24.1%) | <0.001 |
| Dyslipidemia | 4235 (50.8%) | 1413 (59.7%) | 2822 (47.3%) | <0.001 |
| Family history of CAD | 2069 (24.8%) | 374 (15.8%) | 1695 (28.4%) | <0.001 |
| Current smoker | 1076 (12.9%) | 188 (7.9%) | 888 (14.9%) | <0.001 |

### Validation of synthetic CTAC image generation

Likert scale visual analysis on a set of 300 images (including 100 gated, ungated and synthetic CT attenuations scans) revealed Bang’s Blinding Index BI for gated images of 0.58 (95% CI: 0.42–0.74), indicating substantial recognition of gated images. For ungated images, the BI was 0.79 (95% CI: 0.70–0.87), indicating strong recognition of ungated appearance. Only three equivocal ratings (1%) were recorded overall. Gated images were predominantly rated as gated-like (79%), whereas ungated images were primarily rated as ungated-like (79%) but exhibited greater variability. Synthetic ungated images were almost uniformly rated as ungated or ungated-like (97%), with minimal gated-like or equivocal ratings. (**Supplementary Fig. 1**). Overall, these results suggest that the synthetic ungated images preserve the perceptual characteristics of ungated motion, remaining far closer to real ungated images than to gated images, while exhibiting reduced heterogeneity and fewer ambiguous appearances.

Across all gated CAC and ungated CTAC cases, the synthetic CTAC images closely reproduced the noise characteristics of actual CTAC scans. Noise magnitude agreement was strong, with near-unity noise standard-deviation ratios at the voxel level (median 1.03, IQR 0.93–1.13) and at the patch level in whole-body regions (median 0.94, IQR 0.86–1.05) as well as within a cardiac-focused central region (median 0.95, IQR 0.83–1.05). Patch-based distributional comparison demonstrated limited local differences, with median Kolmogorov– Smirnov statistics of 0.046 (IQR 0.037–0.066) globally and 0.048 (IQR 0.032–0.071) in the cardiac region, and corresponding patch-level Wasserstein distances of 6.36 HU and 6.10 HU respectively. Global HU intensity profiles showed substantial overlap between real and synthetic ungated CTAC volumes, with low Jensen–Shannon divergence (0.08). Stratified intensity analysis indicated stronger agreement for high-attenuation voxels (≥130 HU) than for lower-HU regions, consistent with explicit preservation of calcification-relevant structures during synthesis.

### Assessment of DL CAC measures

Assessment of vessel-specific KAD-CAC scoring took 20.6 s ± 2.8 per scan using a graphics processing unit (GeForce RTX 3090, AMD Ryzen 9 5950X 16-core Processors, 64 GB RAM). Examples of DL and expert segmentations are shown in **Figure 1**.

**Figure 1.**
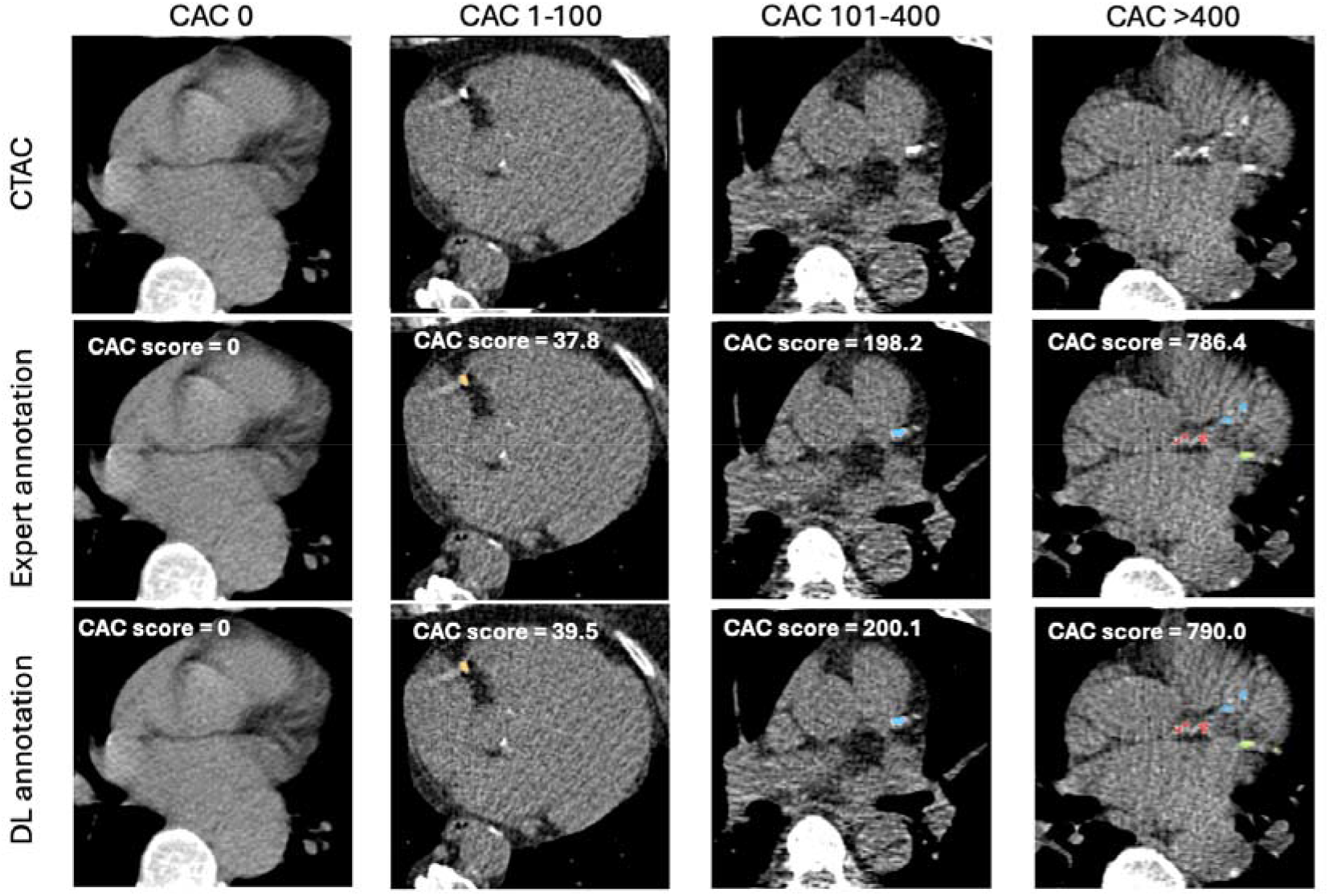
Vessel specific deep learning (DL) coronary artery calcium (CAC) scoring in patients with 0, 1-100, 101-400 and >400 Agatston units (AU) CAC assessed on CT attenuation correction (CTAC) by expert ground truth annotation and DL. Red –left main coronary artery, blue –left anterior descending artery, green –left circumflex artery, yellow-right coronary artery.

The correlation between all DL CAC measures and expert CAC scoring was excellent. There was higher kappa for the KAD-CAC model (Cohen’s kappa 0.86, 95% CI 0.84 – 0.88) compared to the cLSTM model trained on gated scans (Cohen’s kappa K 0.78, 95% CI 0.76 – 0.80), Swin UNETR trained on gated scans (K 0.81, 95% CI 0.80 – 0.82, p<0.001), and Swin UNETR trained on synthetic ungated scans(K 0.84, 95% CI 0.83 – 0.85) with all pairwise differences remaining statistically significant after false discovery rate (FDR) correction (adjusted p < 0.01). Concordance with expert CAC scores was higher for the KAD-CAC model compared to the other DL models in all coronary territories (results in **Supplementary Table 3)**. Per-vessel concordance matrices are shown in **Supplementary Fig. 2-5**. Linear weighted Cohen’s Kappa showed significant improvement in all vessels except LAD when compared to the KAD-CAC model. The results for all models are summarized in **Figure 2**.

**Figure 2.**
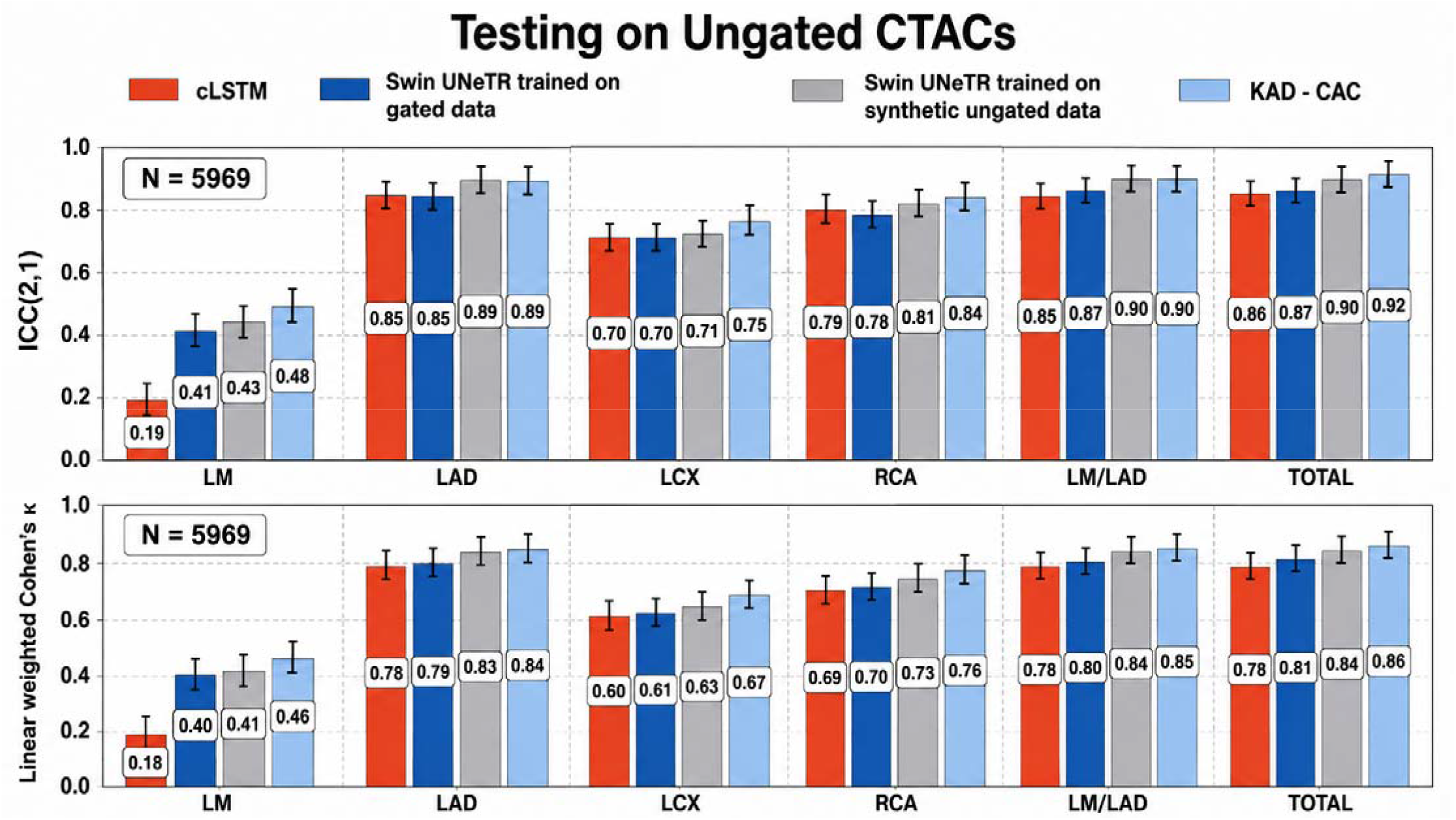
Agreement between DL and expert ground truth. coronary artery calcification (CAC) for the all-vessel score, left main (LM), left anterior descending (LAD), LM/LAD, left circumflex (LCX), and right coronary artery (RCA) for all patients undergoing attenuation correction (AC) CT. p<0.01 for all comparisons to KAD-CAC except for LAD. ICC-intra-class correlation, cLSTM-convolutional long short-term memory network trained on gated data.

For the identification of any CAC on CTAC (identified by expert segmentation), the KAD-CAC model demonstrated excellent diagnostic performance on a global and per-vessel basis. Sensitivity, specificity, negative predictive value (NPV), and positive predictive value (PPV) were 94%, 89%, 95%, and 88% for the LM/LAD territory; 84%, 73%, 71%, and 86% for the LCX territory; and 80%, 84%, 70%, and 90% for the RCA territory, respectively. For total CAC, the KAD-CAC model demonstrated 92% sensitivity, 87% specificity, 88% NPV, and 91% PPV.

### Associations with outcomes

During a median follow-up of 813 days (IQR 498 - 1185), 450 patients experienced either death (total n=349) or MI (total n=122). Incidence of death or MI as a function of CAC category using the KAD-CAC model is shown in **Supplementary Fig. 6.** Compared to patients with CAC 0, patients with CAC 1-100 (unadjusted hazard ratio [HR] 2.56, 95% CI 1.90 – 3.46, p<0.001), CAC 101-400 (unadjusted HR 3.61, 95% CI 2.66 – 4.90, p<0.001) and CAC >400 (unadjusted HR 5.38, 95% CI 4.08 – 7.10, p<0.001) were at increased risk of death or MI. The risk associated with CAC categories in unadjusted and adjusted analyses for each method of CAC scoring are shown in **Table 2**. Kaplan-Meier curves stratified by expert segmentation CAC categories are shown in **Supplementary Fig. 7.** Classification of CAC 0 was discordant between KAD-CAC and expert segmentation in a subset of patients. In adjusted analysis, patients classified as CAC 0 by KAD-CAC had lower risk of death or MI than patients classified as CAC 0 by expert segmentation (adjusted HR 0.50, 95% CI 0.25 – 0.98); conversely, expert CAC 0 carried higher risk relative to KAD-CAC CAC 0 (adjusted HR 2.01, 95% CI 1.02 – 3.97). CAC 0 by the gated-trained (adjusted HR 0.62, 95% CI 0.36 – 1.09), synthetic ungated-trained (adjusted HR 0.59, 95% CI 0.32 – 1.09) and cLSTM (adjusted HR 0.84, 95% CI 0.45 – 1.56) models did not differ significantly from expert CAC 0 (**Figure 3**).

**Figure 3.**
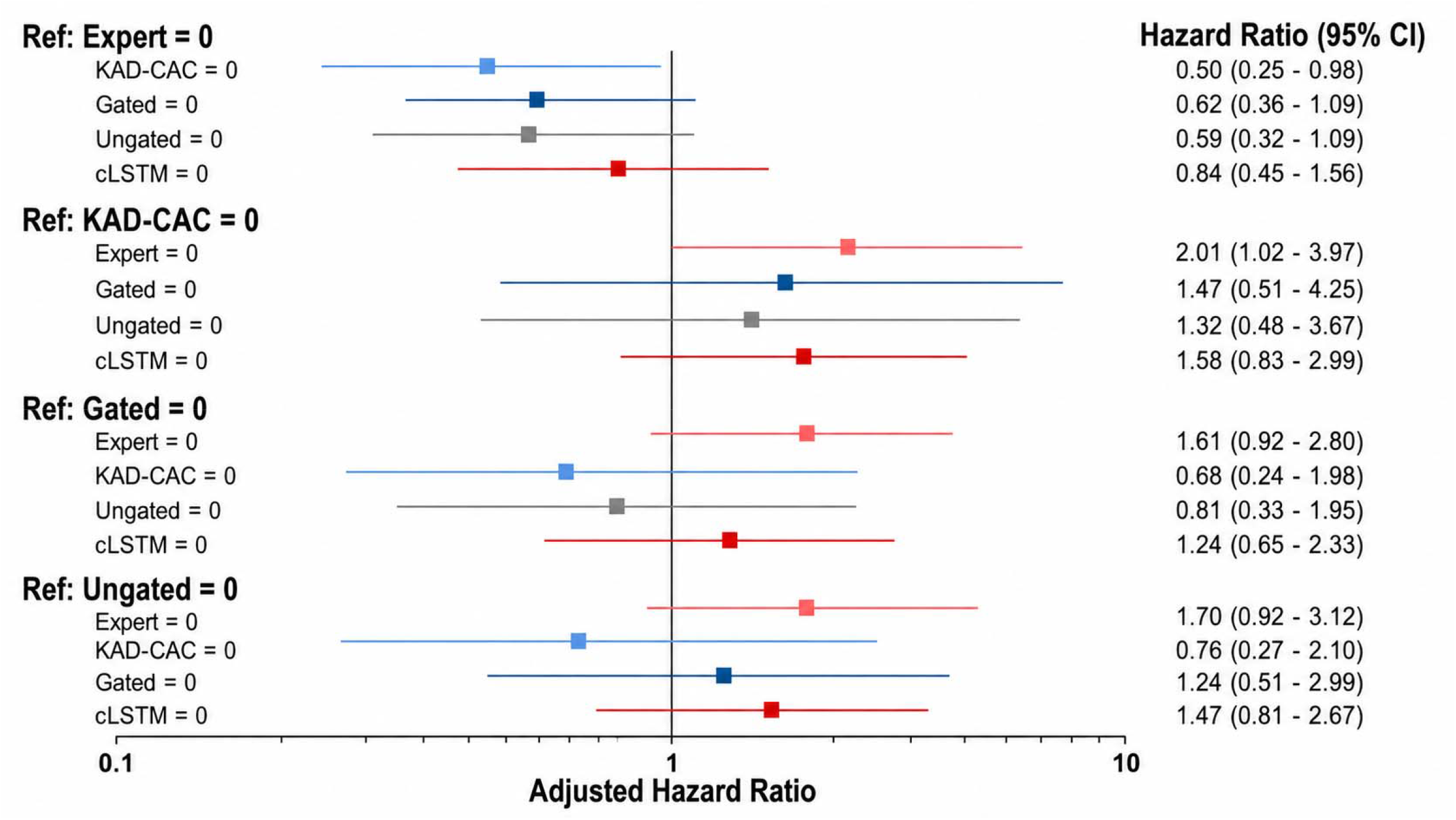
Risk associated with discordant classification of coronary artery calcium score of zero. Patients with CAC of 0 by the KAD-CAC model had significantly lower risk of death or MI compared to patients classified as CAC 0 by expert interpretation.

**Table 2:** Associations with death or myocardial infarction. The multivariable model included age, sex, medical history (hypertension, diabetes, dyslipidemia), family history of CAD, smoking, stress total perfusion deficit and left ventricular ejection fraction. CAC – coronary artery calcium, HR – hazard ratio, LR – likelihood ratio.

|  |  | **Unadjusted HR**<br>**(95% CI)** | **p-value** | **Adjusted HR**<br>**(95% CI)** | **p-value** |
| --- | --- | --- | --- | --- | --- |
| **Expert** | CAC 0 | Reference |  | Reference |  |
| Increase in LR<br>$\chi^2$: 38.5 | CAC 1-100 | 2.04 (1.53 – 2.73) | <0.001 | 1.61 (1.20 – 2.18) | 0.002 |
|  | CAC 101-400 | 3.00 (2.25 – 4.00) | <0.001 | 1.98 (1.45 – 2.69) | <0.001 |
|  | CAC>400 | 4.36 (3.36 – 5.66) | <0.001 | 2.47 (1.83 – 3.28) | <0.001 |
| **KAD-CAC** | CAC 0 | Reference |  | Reference |  |
| Increase in LR<br>$\chi^2$: 60.1 | CAC 1-100 | 2.56 (1.90 – 3.46) | <0.001 | 2.11 (1.56 – 2.87) | <0.001 |
|  | CAC 101-400 | 3.61 (2.66 – 4.90) | <0.001 | 2.54 (1.84 – 3.51) | <0.001 |
|  | CAC>400 | 5.38 (4.08 – 7.10) | <0.001 | 3.14 (2.30 – 4.28) | <0.001 |
| **Swin UNETR**<br>**trained on**<br>**synthetic**<br>**ungated data** | CAC 0 | Reference |  | Reference |  |
| Increase in LR<br>$\chi^2$: 58.1 | CAC 1-100 | 2.41 (1.80 – 3.24) | <0.001 | 1.99 (1.47 – 2.68) | <0.001 |
|  | CAC 101-400 | 3.29 (2.44 – 4.43) | <0.001 | 2.30 (1.68 – 3.14) | <0.001 |
|  | CAC>400 | 5.16 (3.96 – 6.72) | <0.001 | 3.03 (2.25 – 4.09) | <0.001 |
| **Swin UNETR**<br>**trained on gated**<br>**data** | CAC 0 | Reference |  | Reference |  |
| Increase in LR<br>$\chi^2$: 53.7 | CAC 1-100 | 2.42 (1.82 – 3.22) | <0.001 | 1.92 (1.43 – 2.57) | <0.001 |
|  | CAC 101-400 | 3.11 (2.32 – 4.17) | <0.001 | 2.19 (1.61 – 2.98) | <0.001 |
|  | CAC>400 | 4.78 (3.70 – 6.17) | <0.001 | 2.78 (2.09 – 3.72) | <0.001 |
| **cLSTM** | CAC 0 | Reference |  | Reference |  |
| Increase in LR<br>$\chi^2$: 50.1 | CAC 1-100 | 2.15 (1.59 – 2.91) | <0.001 | 1.86 (1.37 – 2.52) | <0.001 |
|  | CAC 101-400 | 3.40 (2.50 – 4.62) | <0.001 | 2.32 (1.68 – 3.20) | <0.001 |
|  | CAC>400 | 4.96 (3.73 – 6.58) | <0.001 | 2.89 (2.11 – 3.95) | <0.001 |

KAD-CAC reclassified risk of death or MI better than expert reading of the same CTAC scans **(Figure 4).** Categorical NRI was highest for the KAD-CAC model (categorical NRI 0.157, 95% CI 0.103 – 0.210) over a baseline model including age, sex, hypertension, diabetes, dyslipidemia, family history, smoking, stress total perfusion deficit, and left ventricular ejection fraction. The positive NRI was 0.051 (95% CI 0.001 to 0.102) and the negative NRI was 0.106 (95% CI 0.088 to 0.123). The categorical NRI for expert CAC annotated on CTACs was compared with the KAD-CAC model using a paired bootstrap (5,000 samples) to account for correlation. The difference in categorical NRI was 0.089 (95% CI 0.051 to 0.125; p <0.001). The difference in categorical NRI was also significant when compared to the model trained with synthetic ungated data (p=0.004) and gated data (p=0.012). Associations with MACE are shown in **Supplementary Table 4 and Supplementary Fig. 8.**

**Figure 4.**
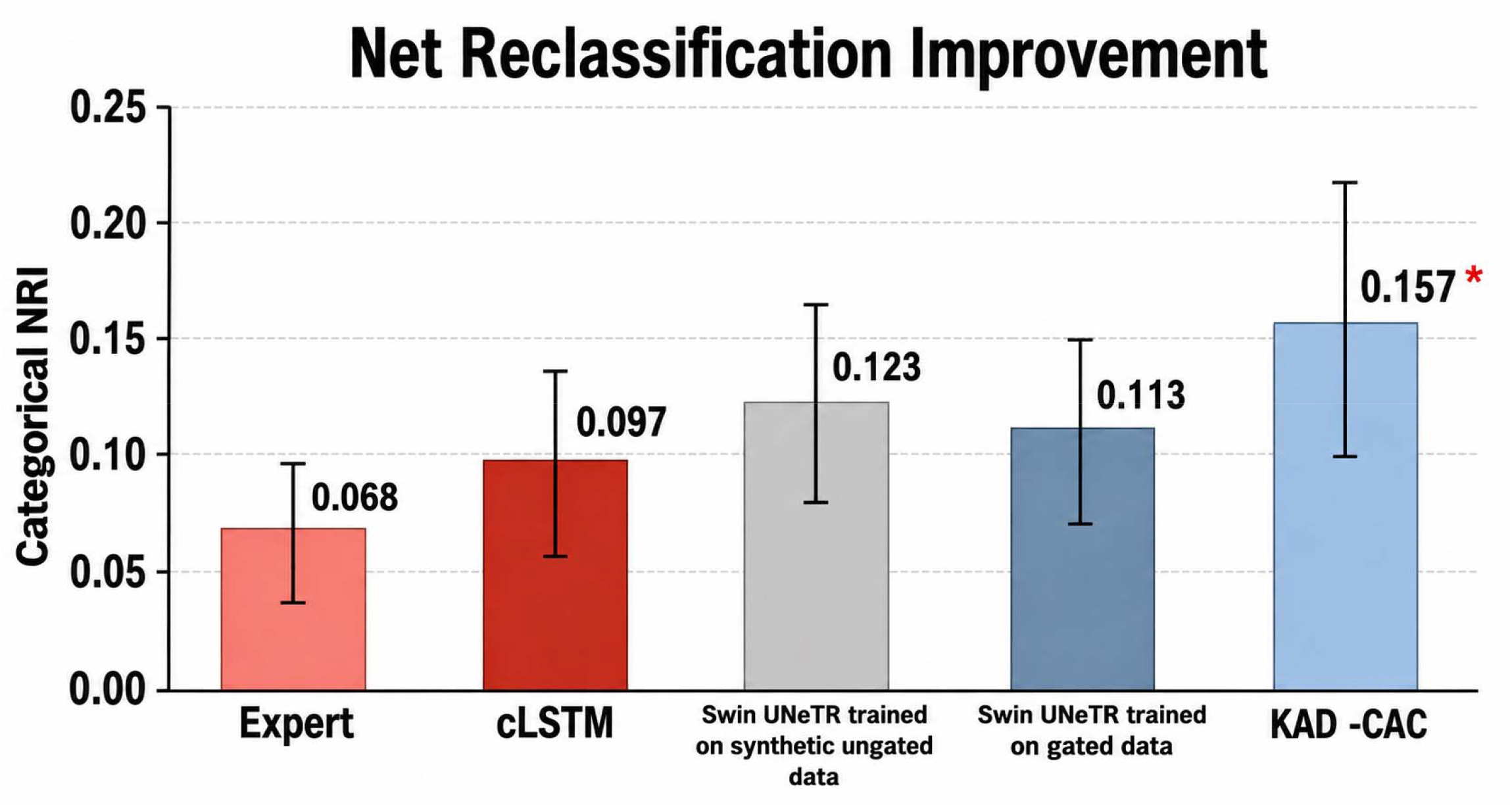
Categorical Net-reclassification improvement (NRI) for CAC through different methods with respect to prediction of death or myocardial infarction. The baseline model included age, sex, hypertension, diabetes, dyslipidemia, family history, smoking, stress TPD, and LVEF as covariates * - p<0.05 compared to all other models. Expert CAC -scoring performed on ungated data. cLSTM model trained on gated data only

### Case Examples

**Supplementary Fig. 9** shows improvement in the LM identification on CTAC imaging from a female patient in her 80s with 3 vessel involvement. The expert LM CAC score was 41 and KAD-CAC LM score was 49. The cLSTM model assigned the LM lesion to the LAD and LCx territories, with LM score of 0. Additional case examples are shown in **Supplementary Fig. 10** and **Supplementary Fig. 11.** Graphical abstract is included in **Figure 5**.

**Figure 5.**
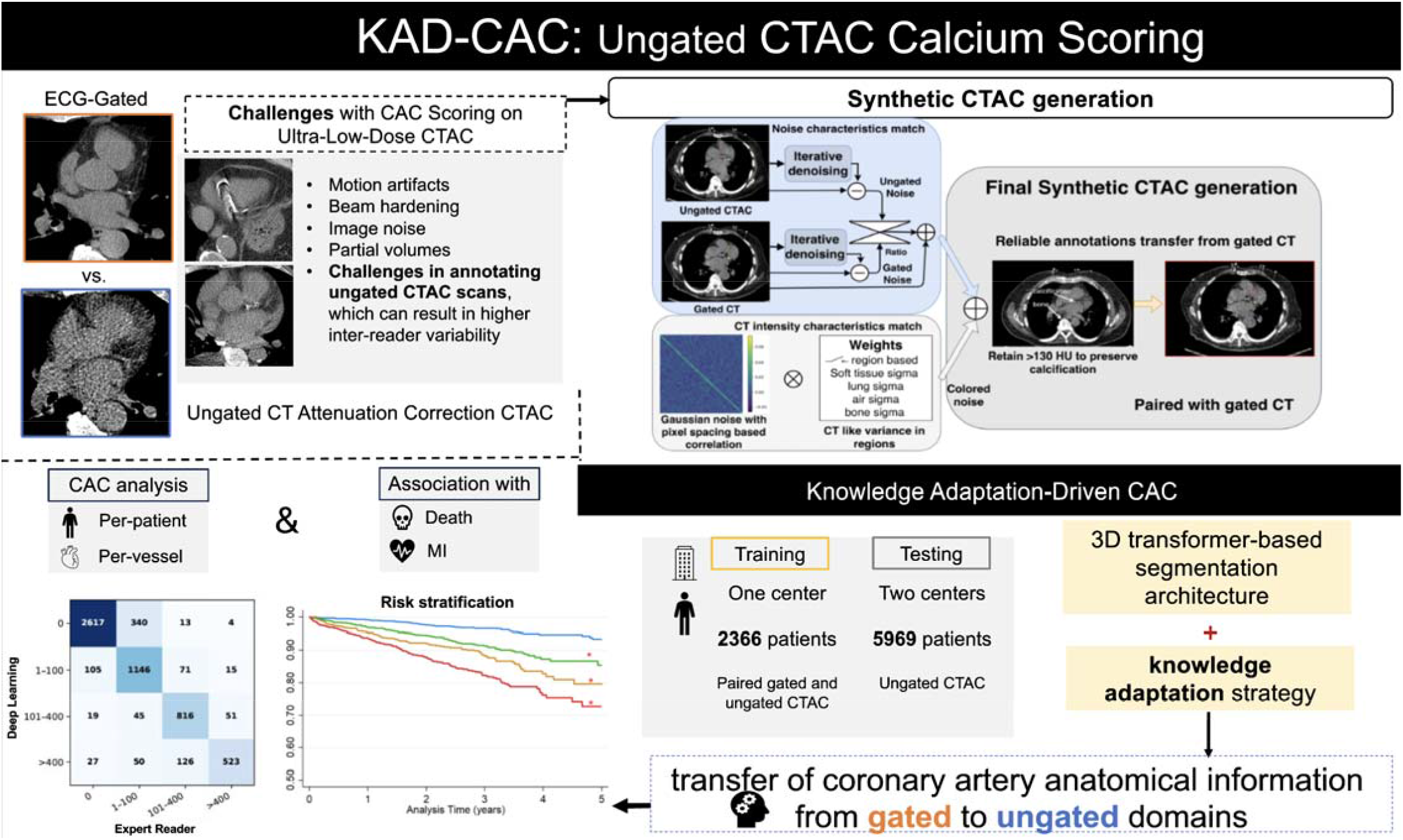
Graphical abstract. Training on synthetic ungated CT derived from expert-annotated gated CT transfers high-quality labels to the ungated domain, improving vessel-specific coronary artery calcium (CAC) scoring on attenuation-corrected CT (CTAC) and reclassification of death or myocardial infarction compared with expert reading in a large, multicenter external population.

## Discussion

The principal finding of this study is that a DL model trained without any labels from CTAC scans predicted death or MI better than expert reading of the same scans in 5969 externally tested patients. To achieve this, we addressed the supervision problem that limits DL on low-dose ungated CT, namely that reference labels are drawn on the same degraded images the model must interpret. This study demonstrated in a large, multi-center external dataset that the resulting model assesses vessel-specific CAC rapidly and accurately, providing improved risk stratification compared to expert segmentations. The central element of the approach is not only domain alignment but label provenance: by synthesizing the ungated image from the gated scan rather than acquiring it separately, every expert annotation made at gated image quality remains voxel-exact on its synthetic ungated counterpart. This gives a lesion-level reference standard that cannot be obtained from real ungated acquisitions. Overall, the synthetic ungated images preserve the perceptual characteristics of ungated motion, remaining far closer to real ungated images than to gated images, while exhibiting reduced heterogeneity and fewer ambiguous appearances.

By training with synthetic CTAC images the model benefits from both expert segmentations from high-quality gated studies to utilize as the ground truth while also learning the image noise and motion artifact patterns of CTAC imaging. KAD-CAC achieved strong-to-excellent agreement with expert measurements for both total and vessel-specific CAC and performed better than networks lacking knowledge adaptation or trained only in the gated or synthetic-ungated domain. DL CAC, by all methods, was an independent predictor of death or MI. Furthermore, the risk reclassification was superior for the proposed KAD-CAC model compared to the other DL models or expert segmentations on ungated CTACs when evaluated in an external population. The proposed KAD-CAC model could provide automated CAC scores in every myocardial perfusion imaging report, without additional imaging or radiation, with prognostic performance at least equal to expert reading of the same scans.

Automated approaches for CAC segmentation have the potential to dramatically improve cardiac risk stratification (2) and guide therapeutic decisions (13, 14). While this information can be obtained from dedicated studies, many chest CT scans performed for other reason could be leveraged for this information opportunistically (7–10). These studies are typically performed without cardiac gating, and with lower radiation exposure (resulting in increased image noise), which can make it challenging to identify small lesions (15). To ensure these limitations were reflected in the training cases, we developed synthetic CTAC images which included both motion artifact and increased image noise (with similar signal to noise as actual ungated imaging). The synthetic images had noise characteristics and an overall appearance that closely approximated actual ungated imaging. The main advantage of using synthetic CTAC images is that they are perfectly registered with the higher-quality, gated CT images (which can then be used as the ground truth segmentation). Previous studies have demonstrated that CAC scores from dedicated ECG-gated CT Agatston have lower interobserver variability compared to CAC scores from low-dose, ungated CT imaging (16). Even expert physicians may struggle with differentiating some CAC lesions from noise on CTAC imaging; whereas this task is relatively simple from dedicated gated imaging. As a result of this training approach, KAD-CAC may identify some CAC lesions which are only readily apparent on dedicated CT scans but are difficult to identify on CTAC imaging alone.

Simulating a degraded acquisition from a higher-quality scan and reusing its labels is an established strategy for low-dose CT denoising and has been applied to organ segmentation when paired low-dose data were unavailable (17–19). In those settings the simulation substitutes for missing paired data, and the transferred labels were themselves generated by a model trained on standard-dose images rather than by experts; whether inherited labels outperform annotation on the degraded image was not examined. The present problem differs in that the reference standard in the target domain is itself less reliable at the lesions of interest: on the same scans, inter-reader agreement for CAC categories is lower on CTAC than on gated CT (kappa 0.85 versus 0.92), the mean absolute inter-reader difference is more than three times larger (41.8 versus 12.2 Agatston units), and the disagreement concentrates at CAC scores below 100 and in the LCx and RCA (20). Deriving the training image from the gated scan therefore supplies supervision of higher fidelity than any annotation obtainable on CTAC, and the degradation model must reproduce ungated cardiac motion in addition to photon noise. To our knowledge, this is the first demonstration that supervision inherited in this way improves patient-level risk prediction beyond the expert reference standard. The same principle applies wherever expert annotation on the acquired image is less reproducible than on a cleaner surrogate, including PET CTAC, low-dose lung screening CT, and accelerated or low-field MRI, although each application requires its own degradation model and external testing. This technical advancement of the KAD-CAC model, and external testing in a large population from two sites, suggest that the model should be generalizable to a large variety of ungated CTAC scans.

While many previous studies have focused solely on total CAC score as the measure of agreement, we also evaluated results on a per-vessel level. This agreement helps ensure that the CAC identified by the DL is the same CAC lesion identified by expert segmentation. However, to date this agreement has only been evaluated in a few studies (21–24). Winkel *et al* tested a convolutional neural network on 262 ECG-gated CT scans and showed a 94% accuracy for the identification of the correct branch, but vessel specific CAC scores were not assessed (25). In the present study, we demonstrated that the KAD-CAC model demonstrated superior per-vessel agreement with expert segmentations across all vascular territories compared to the other DL models. The margin was largest in the left main, the territory in which lesions are smallest and expert annotation on ungated imaging is least reproducible, and therefore the territory in which conventional ungated supervision is weakest. This is the pattern expected if the improvement derives from the fidelity of the training labels rather than from network capacity alone. While this agreement helps support the overall accuracy of the model’s segmentations, it may also provide additional prognostic insights. In non-gated lung cancer screening CT de Vos et al used location specific CAC, along with self-reported cardiovascular risk factors, to predict cardiovascular disease mortality with an AUC of 0.76 (26). In previous work (24), CAC>400 in the left main was associated with the highest risk of MACE. Accordingly, vessel-level accuracy may represent an important step toward clinically actionable, lesion-specific CAC assessment.

The true test for any imaging marker is whether it can provide robust risk stratification to guide therapeutic decisions (27). In this regard, we evaluated associations with death or MI. As expected, we identified a higher risk of death or MI with increasing CAC score for all DL models and expert segmentation. While the differences in risk stratification were small, the KAD-CAC model had the greatest improvement in model fit. Additionally, categorical NRI was significantly higher for the KAD-CAC model compared to expert CAC segmentation as well as compared to the other DL methods. As mentioned above, the unique training approach for KAD-CAC allows it to benefit from expert annotations from dedicated gated images. As a result, and one potential reason for improved risk stratification, KAD-CAC may be able to differentiate small CAC lesions from noise on CTAC imaging. In support of this possibility is the finding that risk in patients classified as CAC 0 by the KAD-CAC model (but not by expert segmentation) was lower compared to patients classified as CAC 0 by expert segmentation (but not by the KAD-CAC model). These results suggest that classification of CAC 0 by the KAD-CAC model could provide reassurance for patients where there is uncertainty regarding cardiovascular risk. In practice, a CAC 0 result from CTAC is the finding most likely to defer preventive therapy, so a method in which patients classified as CAC 0 are at lower risk than the expert equivalent has direct bearing on downstream treatment decisions and subsequent patient outcomes.

This study has a few important limitations. This was a retrospective analysis and future prospective validation is needed. We utilized expert segmentation of CTAC images as the reference standard when assessing agreement with DL scores. However, there remains some uncertainty regarding our ability to accurately identify CAC on CTAC scans. In particular, due to image noise and cardiac motion there will be voxels with inherent uncertainty in classification. For the same reason, we could not include a conventionally supervised comparator — a Swin UNETR trained on real CTAC images with expert CTAC labels — which would have permitted a direct contrast between synthetic paired training and standard supervision in the target domain. Expert CTAC annotations were available only in the Yale and Calgary cohorts, which were reserved for external testing and were not used in any way for model development; training on them would have forfeited the held-out nature of the evaluation. Such a comparator would also inherit the reader uncertainty it was intended to benchmark, since inter-reader agreement for CAC categories is lower on CTAC than on gated CT (kappa 0.85 versus 0.92) (20). We excluded patients with known coronary artery disease including previous revascularization and therefore did not evaluate how well the DL models differentiate stents or surgical clips from CAC. However, it is unlikely that the extent of CAC would significantly shift the cardiovascular risk of those patients since they would all typically be considered high risk.

We developed a transformer-based segmentation architecture with cross domain knowledge transfer strategy for CAC segmentation. More specifically, we generated synthetic ungated images from paired gated CAC scans so that the model learns from both perfectly co-registered high-quality expert segmentations and the image noise and motion characteristics of CTAC imaging. The KAD-CAC model provided better agreement with expert annotations compared to DL models trained without synthetic CTAC images or models trained using either the gated or ungated images alone. Furthermore, this approach provides superior risk stratification for death or MI compared to expert segmentation or other DL approaches.

## Methods

### Study Population

In this multi-center study, we used imaging and clinical data from three centers to train and test a knowledge adaptation-based approach for quantifying vessel specific CAC scores. This included internal training (n=2058) and validation (n=308) on ECG gated non-contrast cardiac CT from Cedars-Sinai Medical Centre (Los Angeles, US). External testing was performed using low-dose, ungated CTAC imaging (n=5969) from patients undergoing SPECT/CT myocardial perfusion imaging at either Yale (New Haven, US) or the University of Calgary (Calgary, Canada) (28). Only the first study for each patient was included in the analysis. Patients with previous myocardial infarction (MI), percutaneous coronary intervention or coronary artery bypass grafting were excluded. This study was approved by the institutional review board at each site and was performed in accordance with the Declaration of Helsinki. Depending on local requirements, written informed consent was obtained or the requirement was waived for analysis of deidentified data

### CT Image acquisition and image analysis

Detailed information regarding image acquisition in the study populations has been published previously (8, 29, 30). Additional details are available in the supplement.

Expert quantification of CAC was performed using dedicated software (QPET Suite, Cedars Sinai Medical Centre, Los Angeles). CAC scores were calculated on a per-vessel basis using the Agatston method for both ECG gated CT in training and CT AC imaging in test(31). The ground truth expert derived CAC scores were segmented by experienced technologists and clinicians with >10 years’ experience, following previously described vessel-territory annotation criteria (24).

### Deep learning model architecture and training

An end-to-end DL framework was developed to enable accurate CAC quantification from synthetic ungated CT attenuation correction (CTAC) scans by leveraging anatomical knowledge from gated CAC scans. The proposed method (KAD-CAC) uses a 3D transformer-based segmentation architecture with a cross-domain knowledge transfer strategy to transfer coronary artery anatomical information from gated to ungated domains(32). The overall approach is designed to mimic expert reader behavior by exploiting three-dimensional spatial context across contiguous CT slices.

### Network Architecture

The proposed framework follows the architecture shown in **Figure 6**. Additional details are available in the supplement. To ensure that the knowledge adaptation from gated scans benefits the overall model training, we compared against training the 3D Swin UNeTR with 1) gated data only and 2) synthetic ungated data only (**Supplemental Figure 12**). These comparator models were selected to sequentially evaluate the contributions of model architecture, synthetic ungated training, and knowledge adaptation. Specifically, comparisons between cLSTM and gated Swin UNETR assessed the effect of the 3D transformer architecture, gated and synthetic-ungated Swin UNETR assessed the effect of synthetic-domain training, and synthetic-ungated Swin UNETR and KAD-CAC assessed the additional contribution of cross-domain knowledge adaptation.

**Figure 6.**
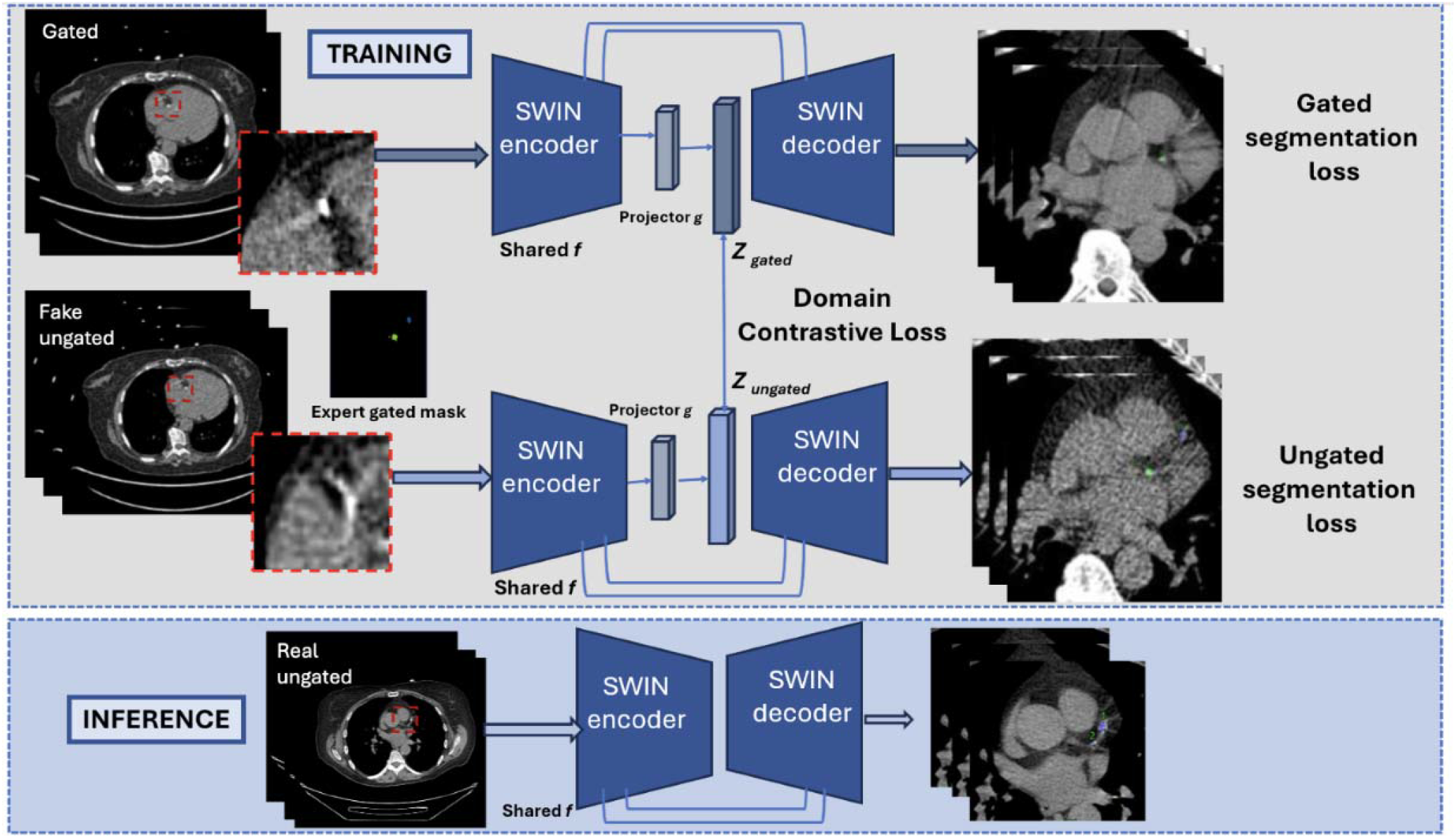
Overview of model training and testing. The model was trained using a cohort with paired gated and ungated computed tomography (CT) scans via a knowledge adaptation strategy. We developed a synthetic ungated CT, with the model trained to segment CAC from synthetic ungated imaging with expert segmentations from gated chest CT as the ground truth. End-to-end model includes cost function as a weighted sum of gated segmentation loss, ungated segmentation loss and domain aware contrastive loss from embeddings *Z_gated,_ Z_ungated_* obtained through a linear projector *g.* The network encoder *f* is shared during training. The red box indicates a zoom into the region as sliding window-based input and output volumes are generated.

### Creation of Synthetic Ungated CTAC Volumes

Direct voxel-wise pairing between separately acquired gated CAC scans and ungated CTAC scans is challenging due to respiratory and cardiac motion, differences in breath-hold status, and partial volume effects(20), which can lead to misclassification of CAC (**Supplementary Fig. 13**) showing label uncertainty. To address this limitation, paired synthetic ungated CTAC volumes were generated from gated CAC scans using a noise model derived from real ungated CTAC data. Because the synthetic volume is derived from the gated scan itself rather than acquired separately, the gated expert segmentation remains valid voxel-for-voxel on the synthetic ungated image without any registration step. Each training example therefore pairs an image carrying CTAC-like noise and motion with a mask annotated at gated image quality — a combination that cannot be obtained from any real pair of acquisitions (**Figure 7**). Expert segmentation of ECG-gated CT scans has improved reproducibility compared to ungated, low-dose CTAC imaging (20). A blinded evaluation of images was used to evaluate the visual comparability of synthetic ungated imaging to actual ungated imaging. Additional details are available in the supplement.

**Figure 7.**
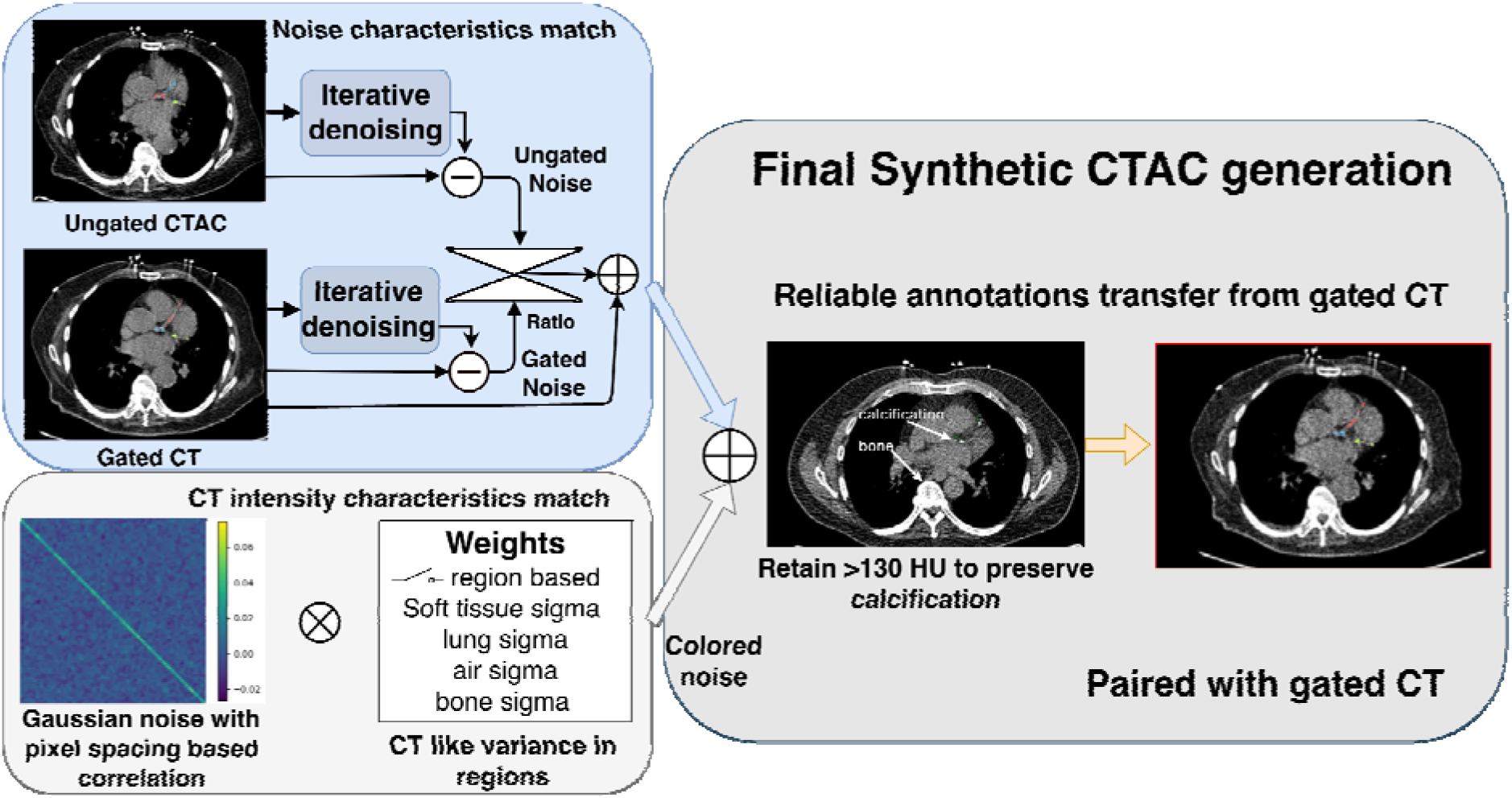
Synthetic CTAC creation pipeline: Ungated CTAC and gated CT scan pairs used to estimate noise using alternating direction method of multipliers (ADMM) -based penalized weighted least squares total variation (PWLS-TV). Noise statistics from ungated CTAC scans were then computed and injected into the gated volumes, with a scaling factor applied to match the standard deviation observed in real ungated data for noise characteristics to match. For CT intensity characteristics to match, we augmented the data with CT-like Gaussian noise rather than simple white noise. White noise was first spatially correlated using a Gaussian filter to match the characteristic grain size imposed by CT reconstruction kernels and slice thickness. The noise amplitude was then modulated as a function of Hounsfield Units HU, with higher variance in air and lung regions and lower variance in soft tissue and bone, reflecting the physics of CT image formation through colored noise. Voxels corresponding to calcified lesions (≥130 HU) were preserved to prevent artificial degradation of CAC regions. Sigma defines the standard deviation in different sections.

### External testing

The DL model was tested on external ungated data from two sites which were not used in any way for model training. Per-vessel CAC scores for the expert ground truth on the ungated CTACs and DL models were categorized into four groups (0, 1-100, 101-400 and >400 Agatston units [AU]).

### Clinical outcomes

Beyond agreement with expert CAC measurements, prognostic analyses examined the composite of death or myocardial infarction (MI). Our secondary outcome was incidence of major adverse cardiovascular events (MACE), which included death, MI, admission for unstable angina, or revascularization. Incidence of death was ascertained through the national death index in patients from Yale University and Alberta Vital statistics at the University of Calgary. Incidence of MI and admission for unstable angina was adjudicated by experienced clinicians at each site using standard criteria (32).

### Statistical Analysis

Details are available in the supplement.

## Data Availability

To the extent allowed by data sharing agreements and institutional review board protocols, data and codes used in this manuscript will be shared upon written request.

## Code Availability

cLSTM code is publicly available under a Creative Commons BY-NC license at https://doi.org/10.5281/zenodo.10632288. SWIN-UNETR code available: https://monai.readthedocs.io/en/0.9.1/_modules/monai/networks/nets/swin_unetr.html

## Acknowledgements

This research was supported by grant R35HL161195 from the National Heart, Lung, and Blood Institute/ National Institutes of Health (NHLBI/NIH) (PI: Piotr Slomka). The content is solely the responsibility of the authors and does not necessarily represent the official views of the National Institutes of Health.

## Author contributions

AS, □RJHM, and □RL, □PJS conceptualized and designed this study. AMM, AS, □ASK □ML, RJHM, and PJS participated in the preparation of the study’s data materials. RJHM, DB, AF, EM□participated□in data acquisition. AS,□and□AK□designed and implemented the deep learning pipeline. AS and□ML□accessed and verified the underlying data reported in the manuscript. AS,□RJH□and □RL, □PJS drafted the manuscript, which was critically reviewed and evaluated by all other co-authors. Drs. Leahy and Slomka contributed equally to this work and are joint senior authors. All authors□read and approved the final manuscript and had final responsibility for the decision to□submit□for publication. The institutional review board at each participating site approve the study protocol, which complied with the Declaration of Helsinki.

## Competing interests

Dr. Robert Miller has received research support from Alberta Innovates and consulting fees from Alnylam and Bayer. Drs. Berman and Slomka and Mr. Kavanagh participate in software royalties for QPS software at Cedars-Sinai Medical Center. Dr. Marcinkiewicz received consulting fees from APQ Health. Dr. E. Miller received grant support from Pfizer, ARGO SPECT, Alnylam, Siemens□Medical Systems□and the National Institutes of Health. Dr. E. Miller□received consulting fees from Pfizer, Alnylam, Synektik, and Eidos/BioBridge. Dr. Dey, Dr. Slomka, and Dr. Berman declare equity interest in APQ Health. Prof. Slomka, and Dr. Berman participate in software royalties for QPS software at Cedars-Sinai Medical Center. Dr Berman received research grant support from the Dr. Miriam and Sheldon G. Adelson□Medical Research Foundation, served as a consultant for GE Healthcare. Prof. Slomka received research grant support from Siemens Medical Systems and consulting fees from Synektik S.A. Other authors declare no competing interests.

## Consent

The registry protocols were approved by the Institutional Review Board at all participating sites, and sites either obtained written informed consent or a waiver of consent for the use of the deidentified data. The overall registries were approved by the investigational review board at Cedars-Sinai Medical Center and complied with the Declaration of Helsinki. Clinical variables collected at each site were de-identified and anonymized prior to transfer to the core site using dedicated software compliant with the Health Insurance Portability and Accountability Act (HIPAA).

## Notes

### Competing Interest Statement

Dr. Robert Miller has received consulting fees from Alnylam and Bayer and research support from Alberta Innovates. Drs. Berman and Slomka and Mr. Kavanagh participate in software royalties for QPS software at Cedars-Sinai Medical Center. Dr. Marcinkiewicz received consulting fees from APQ Health. Dr. E. Miller received grant support from Pfizer, ARGO SPECT, Alnylam, Siemens Medical Systems and the National Institutes of Health. Dr. E. Miller received consulting fees from Pfizer, Alnylam, Synektik, and Eidos/BioBridge. Dr. Dey, Dr. Slomka, and Dr. Berman declare equity interest in APQ Health. Prof. Slomka, and Dr. Berman participate in software royalties for QPS software at Cedars-Sinai Medical Center. Dr Berman received research grant support from the Dr. Miriam and Sheldon G. Adelson Medical Research Foundation, served as a consultant for GE Healthcare. Prof. Slomka received research grant support from Siemens Medical Systems and consulting fees from Synektik S.A. Other authors declare no competing interests.

### Summary of Updates

We revised the title, abstract, introduction, and discussion of the manuscript.

