## Supplemental Material for "Deep Learning Trained Without Target-Domain Labels Improves Risk Stratification Over Expert Coronary Calcium Reading on Routine Attenuation Correction CT"

**Supplemental Methods**

CT Image acquisition and image analysis

Dedicated CAC CT at the training center was acquired at end inspiration with prospective ECG triggering (120 kVp, 85-150 mAs, and 3-mm sections). CTAC scans were obtained during free breathing without ECG synchronization. The training-site protocol used 100 kVp, 11-13 mAs, and 5-mm sections. At the first external site (Calgary), images were acquired at 120 kVp and 20 mA with 2.5-mm sections; at the second external site (Yale), the protocol used 120 kVp and a body-mass-index-dependent tube current of 60-150 mA.

Creation of Synthetic Ungated CTAC Volumes

Both gated and ungated CT volumes were first denoised using an ADMM-based penalized weighted least squares total variation approach to estimate their underlying noise characteristics (*1*). Noise statistics from ungated CTAC scans were then computed and injected into the gated volumes, with a scaling factor applied to match the standard deviation observed in real ungated data. To reduce the appearance gap between volumes, we augmented the data with Gaussian noise exhibiting CT-like spatial correlation and intensity-dependent variance, rather than spatially uncorrelated (white) noise. White noise was first spatially correlated using a Gaussian filter to match the characteristic grain size imposed by CT reconstruction kernels and slice thickness. The noise amplitude was then modulated as a function of Hounsfield Units HU, with higher variance in air and lung regions and lower variance in soft tissue and bone, reflecting the physics of CT image formation. This procedure produces noise textures and intensity distributions that visually and quantitatively better match real clinical CT images. Voxels corresponding to calcified lesions (≥130 HU) were preserved to prevent artificial degradation of CAC regions. The resulting synthetic ungated volumes underwent visual inspection by an experienced technologist and were rated using a five-point Likert scale (*2*), demonstrating acceptable realism and similarity to true ungated CTAC scans as well as blinded score index. For quantitative validation, synthetic volumes were spatially resampled to the corresponding ungated CTAC geometry using linear interpolation to ensure alignment for comparison. Noise similarity between real and synthetic ungated CTAC volumes was assessed using both voxel-wise and patch-based analyses. Patch-based evaluation was performed using large three-dimensional patches sampled from whole-body regions and only in the cardiac-focused central region. Agreement was quantified using noise standard-deviation ratios(3), Kolmogorov–Smirnov (KS) statistics (4), and Wasserstein distances as effect-size measures (5). Intensity profile similarity was evaluated using Jensen–Shannon divergence(6) and Wasserstein distance computed on HU histograms (7), with additional stratified analysis below and above 130 HU to separately assess soft-tissue/background regions and high-attenuation structures.

Network Architecture:

A shared 3D Swin Transformer encoder $f\left( . \right)$ processes both gated CAC volumes and ungated (synthetic or real) CTAC volumes. The encoder extracts hierarchical volumetric features capturing long-range coronary anatomy. These shared features are used for two objectives: (1) supervised CAC segmentation via decoder heads and (2) cross-domain knowledge adaptation via contrastive learning.

Let $X_{gated}$ and $X_{ungated}$ denote the gated and ungated input volumes, respectively. Both inputs are passed through the shared encoder:
$H_{gated}=f\left( X_{gated} \right), H_{ungated}=f\left( X_{ungated} \right)$	**(equation 1)**

For knowledge adaptation, the encoder outputs are mapped to a latent embedding space using a projection head **g:**
$Z_{gated}=g\left( H_{gated} \right), Z_{ungated}=g\left( H_{ungated} \right)$	**(equation 2)**

The projection head is used only during training. During inference, only the encoder $f\left( . \right)$ and the ungated decoder are retained.

### *Segmentation Loss: Focal Loss*

CAC segmentation is characterized by extreme class imbalance, with calcified voxels representing a very small fraction of the total volume, particularly in vessels such as the left main (LM) and left circumflex (LCX) arteries. To address this imbalance, focal loss was used for both gated and ungated segmentation outputs (8). For a voxel with ground-truth label **y ∈ {0,1}** and predicted probability **p**, the focal loss is defined as

$L_{focal}\left( p,y \right)=-\alpha*y*\left( 1-p \right)^{\gamma}*\log\left( p \right)-\left( 1-\alpha\right)*\left( 1-y \right)*p^{\gamma}*\log\left( 1-p \right)$ **(equation 3)**

where **α** balances foreground and background contributions and **γ** is the focusing parameter.

The total segmentation loss is computed as:

$L_{seg}=L_{focal}^{gated}+L_{focal}^{ungated}$	**(equation 4)**

### *Knowledge Adaptation Loss: NT-Xent Loss*

To transfer coronary artery anatomical knowledge from gated CAC scans to ungated CTAC scans, a contrastive loss based on the normalized temperature-scaled cross entropy (NT-Xent) formulation was employed(9). This loss encourages latent representations of paired gated and ungated scans from the same patient to be close in the embedding space, while separating representations from different patients.

The cosine similarity between two embeddings $Z_{i}$	and $Z_{j}$	is defined as:

$sim\left( Z_{i},Z_{j} \right)=\left( Z_{i}\cdot Z_{j} \right)/\left( \left| \left| Z_{i} \right| \right|_{2}*\left| \left| Z_{j} \right| \right|_{2} \right)$	**(equation 5)**

For a batch of **N** paired samples *i*, the NT-Xent loss for the positive pair ($Z_{gated}^{i}, Z_{ungated}^{i}$ ) is given by:

$L_{NT-Xent}^{i}=-\log\left( \exp\left( sim\left( Z_{gated}^{i},Z_{ungated}^{i} \right)/\tau\right)/\Sigma_{\left( k=1,k\neq i \right)}^{N}\exp\left( sim\left( Z_{gated}^{i},Z_{ungated}^{k} \right)/\tau\right) \right)$**(equation 6)**

where **τ** is the temperature parameter

The final contrastive loss is obtained by averaging across the batch:

$L_{contrastive}= \left( 1/N \right)*\Sigma_{\left( i=1 \right)}^{N}L_{NT-Xent}^{i}$	**(equation 7)**

### *Total Training Objective*

The complete end-to-end training objective is defined as:

$L_{total}=L_{seg}+\lambda*L_{contrastive}$

**(equation 8)**

where **λ** controls the relative contribution of the knowledge adaptation loss. This joint objective balances voxel-level accuracy with representation-level alignment. The segmentation loss ensures accurate CAC delineation, while the contrastive loss ensures that ungated CTAC scans benefit from the rich anatomical information present in gated CAC scans.

The network was trained using a focal loss with focusing parameter γ = 5. The contrastive loss was weighted by λ = 10 and used a temperature parameter τ = 0.1. Training was performed for a maximum of 500 epochs using the Adam optimizer with an initial learning rate of 5 × 10⁻⁴ and weight decay of 1 × 10⁻⁵. The learning rate was controlled using ReduceLROnPlateau with a reduction factor of 0.1, patience of 15 epochs, and minimum learning rate of 1 × 10⁻⁷. Three-dimensional 64 × 64 × 32-voxel patches were used for training. Data augmentation included random rotation of up to 10 degrees, and translation of 15 pixels, each applied with a probability of 0.5. The Swin UNETR used a feature size of 48 and a 512-dimensional contrastive projection head. Automatic mixed-precision training was disabled, and gradients were clipped to a maximum norm of 10.

The convolutional LSTM (cLSTM) network was trained on gated data only. The cLSTM network works on 2.5 D approach where slice of interest and adjacent slices of interest (3 slices in total) are taken into consideration, whereas the Swin UNETR works on 3D sliding windows with 50% overlap.

Statistical Analysis:

Continuous variables are presented as mean and standard deviation for normally distributed data and median and interquartile range (IQR) for non-normally distributed data. Categorical variables are presented as frequency and percentage. Bang blinding index (BI) for two classes was used to assess the quality of synthetic ungated CT images compared to actual ungated CT images (*10*). Ordinal agreement was assessed using linear weighted Cohen’s kappa and concordance correlation coefficients, as well as intra-class correlations. Correlation coefficients and Cohen’s kappa were interpreted as follows: <0.2 - very weak, 0.2 to <0.40 - weak, 0.40 to <0.60 - moderate, 0.6 to <0.80 - strong, and 0.8 to 1 - excellent. Statistical significance was assessed using the Wilcoxon rank-sum test and Pearson’s Chi-squared test. The per-vessel diagnostic accuracy for the identification of CAC of the DL model compared to the expert ground truth was assessed using sensitivity, specificity, positive predictive value (PPV), and negative predictive value (NPV). Associations with death or MI were assessed using Cox proportional hazards models. Multivariable models included age, sex, hypertension, diabetes, dyslipidemia, family history, smoking, stress TPD, and LVEF as covariates.

To determine the incremental prognostic value of CAC, categorical net reclassification improvement (NRI) was calculated by comparing the baseline multivariable model with and without the addition of CAC categories. Predicted 5-year event risk was classified into four clinically relevant categories: low (<2.5%), low-intermediate (2.5% to <5%), intermediate (5% to <10%), and high (≥10%). The NRI quantifies the extent to which adding CAC correctly reclassifies individuals into higher-risk categories among those who experienced an event and into lower-risk categories among those who remained event-free. Positive NRI values indicate improved risk classification with the addition of CAC. Differences in NRI between CAC scoring methods were assessed using paired bootstrap resampling to account for the correlated nature of predictions derived from the same study population. A two tailed p-value <0.05 was considered statistically significant. For secondary analyses involving multiple correlated pairwise comparisons of agreement metrics, p-values were adjusted using the Benjamini–Hochberg false discovery rate (FDR) procedure (*11*). Analyses were performed using Stata version 13.1 (StataCorp, Texas), R (Version 4.2.3, R Foundation for Statistical Computing, Vienna, Austria), and Python 3.7.4.

|  | Variable | Median (Q1, Q3) |
| --- | --- | --- |
| Expert Segmentation | Total CAC score | 19 (0, 261) |
|  | LM CAC score | 0 (0, 0) |
|  | LCx CAC score | 0 (0, 16) |
|  | LAD CAC score | 5 (0, 122) |
|  | RCA CAC score | 0 (0, 24) |
| Swin UNETR trained on synthetic ungated data | Total CAC score | 19 (0, 257) |
|  | LM CAC score | 0 (0, 0) |
|  | LCx CAC score | 0 (0, 6) |
|  | LAD CAC score | 0 (0, 127) |
|  | RCA CAC score | 0 (0, 31) |
| Swin UNETR trained on gated data | Total CAC score | 14 (0, 253) |
|  | LM CAC score | 0 (0, 0) |
|  | LCx CAC score | 0 (0, 3) |
|  | LAD CAC score | 0 (0, 118) |
|  | RCA CAC score | 0 (0, 26) |
| KAD-CAC | Total CAC score | 21 (0, 264) |
|  | LM CAC score | 0 (0, 0) |
|  | LCx CAC score | 2 (0, 10) |
|  | LAD CAC score | 0 (0, 130) |
|  | RCA CAC score | 0 (0, 33) |
| cLSTM | Total CAC score | 25 (0, 256) |
|  | LM CAC score | 0 (0, 0) |
|  | LCx CAC score | 0 (0, 14) |
|  | LAD CAC score | 0 (0, 114) |
|  | RCA CAC score | 0 (0, 43) |

**Supplemental Table 1.** Total and per-vessel coronary artery calcium (CAC) scores for each of the models. cLSTM- convolutional long, short term memory network. Q1- quartile1, Q3- quartile 3

| Cohort | Variable | CAC score, Median (Q1, Q3) | Total lesions | Patients with ≥1 lesion |
| --- | --- | --- | --- | --- |
| Training (N = 2,058) | Total | 94 (0, 464) | 11,902 | 1,221 |
|  | LM | 0 (0, 0) | 503 | 382 |
|  | LCx | 0 (0, 55) | 3,222 | 916 |
|  | LAD | 42 (0, 240) | 5,417 | 1,338 |
|  | RCA | 0 (0, 79) | 4,785 | 945 |
| Validation (N = 308) | Total | 104 (5, 494) | 1,971 | 193 |
|  | LM | 0 (0, 0) | 77 | 62 |
|  | LCx | 0 (0, 66) | 539 | 138 |
|  | LAD | 51 (0, 261) | 909 | 210 |
|  | RCA | 2 (0, 95) | 879 | 155 |

**Supplemental Table 2.** Total and per-vessel coronary artery calcium (CAC) scores in training and internal validation through expert annotations. Q1- quartile1, Q3- quartile 3

|  |  | Kappa (95% CI) | Agreement (95% CI) |
| --- | --- | --- | --- |
| Total | cLSTM | 0.78 (0.76 – 0.80) | 0.76 (0.75 - 0.78) |
|  | KAD-CAC | 0.86 (0.84 - 0.88) | 0.84 (0.83 - 0.85) |
|  | Gated data trained Swin UNETR | 0.81 (0.80 - 0.82) | 0.80 (0.79 - 0.81) |
|  | Synthetic Ungated data trained Swin UNETR | 0.84 (0.83 - 0.85) | 0.82 (0.81 - 0.83) |
| LAD | cLSTM | 0.78 (0.77 - 0.79) | 0.80 (0.78 - 0.81) |
|  | KAD-CAC | 0.84 (0.84 - 0.85) | 0.85 (0.85 - 0.86) |
|  | Gated data trained Swin UNETR | 0.79 (0.78 - 0.80) | 0.81 (0.80 - 0.82) |
|  | Synthetic Ungated data trained Swin UNETR | 0.83 (0.83 - 0.84) | 0.84 (0.83 - 0.85) |
| LCx | cLSTM | 0.60 (0.58 - 0.62) | 0.74 (0.73 - 0.76) |
|  | KAD-CAC | 0.67 (0.66 - 0.68) | 0.80 (0.79 - 0.81) |
|  | Gated data trained Swin UNETR | 0.61 (0.59 - 0.63) | 0.78 (0.77 - 0.79) |
|  | Synthetic Ungated data trained Swin UNETR | 0.63 (0.61 - 0.65) | 0.78 (0.77 - 0.79) |
| LM | cLSTM | 0.18 (0.16 - 0.21) | 0.82 (0.81 - 0.83) |
|  | KAD-CAC | 0.46 (0.44 - 0.48) | 0.85 (0.84 - 0.86) |
|  | Gated data trained Swin UNETR | 0.40 (0.38 - 0.42) | 0.84 (0.83 - 0.85) |
|  | Synthetic Ungated data trained Swin UNETR | 0.41 (0.39 - 0.43) | 0.84 (0.83 - 0.85) |
| RCA | cLSTM | 0.69 (0.67 - 0.71) | 0.76 (0.75 - 0.77) |
|  | KAD-CAC | 0.76 (0.75 - 0.77) | 0.82 (0.81 - 0.83) |
|  | Gated data trained Swin UNETR | 0.70 (0.69 - 0.71) | 0.79 (0.78 - 0.80) |
|  | Synthetic Ungated data trained Swin UNETR | 0.73 (0.72 - 0.74) | 0.80 (0.79 - 0.81) |
| LM/LAD | cLSTM | 0.78 (0.77 - 0.79) | 0.76 (0.75 - 0.77) |
|  | KAD-CAC | 0.85 (0.84 - 0.86) | 0.86 (0.85 - 0.87) |
|  | Gated data trained Swin UNETR | 0.80 (0.79 - 0.82) | 0.81 (0.80 - 0.82) |
|  | Synthetic Ungated data trained Swin UNETR | 0.84 (0.83 - 0.85) | 0.85 (0.84 - 0.86) |

**Supplemental Table 3.** Linear weighted kappa and agreement (concordance correlation coefficient) on a per-patient and per-vessel basis.

|  | | | **Univariable analysis** | | **Multi-variable analysis *** | |
| --- | --- | --- | --- | --- | --- | --- |
|  |  |  | **HR (95% CI)** | **P value** | **HR (95% CI)** | **P value** |
| (A) | Total | 1-100 | 2.17 (1.74, 2.7) | <0.001 | 1.95 (1.56, 2.44) | 0.004 |
|  |  | 101-400 | 3.54 (2.85, 4.4) | <0.001 | 2.95 (2.34, 3.72) | <0.001 |
|  |  | >400 | 5.74 (4.72, 6.98) | <0.001 | 4.35 (3.49, 5.43) | <0.001 |
|  | LM/LAD | 1-100 | 2.47 (2.04, 2.99) | <0.001 | 2.07 (1.69, 2.52) | <0.001 |
|  |  | 101-400 | 3.59 (2.94, 4.37) | <0.001 | 2.73(2.2, 3.38) | <0.001 |
|  |  | >400 | 5.26 (4.31, 6.43) | <0.001 | 3.75 (2.99, 4.69) | <0.001 |
|  | LCX | 1-100 | 2.25 (1.92, 2.64) | <0.001 | 1.8 (1.52, 2.13) | <0.001 |
|  |  | 101-400 | 3.46 (2.83, 4.23) | <0.001 | 2.54 (2.06, 3.14) | <0.001 |
|  |  | >400 | 4.14 (3.12, 5.49) | <0.001 | 2.85 (2.13, 3.8) | <0.001 |
|  | RCA | 1-100 | 2.07 (1.74, 2.46) | <0.001 | 1.74 (1.45, 2.08) | <0.001 |
|  |  | 101-400 | 3.16 (2.61, 3.82) | <0.001 | 2.41 (1.98, 2.94) | <0.001 |
|  |  | >400 | 4.23 (3.47, 5.17) | <0.001 | 2.91 (2.34, 3.61) | <0.001 |

* Adjusted for age, sex, total perfusion deficit, left ventricular ejection factor, hypertension, diabetes mellitus, dyslipidemia, family history, and smoking status.

Reference group = zero CAC.

CI, confidence interval; LAD, left anterior descending; LCX, left circumflex; LM, left main; HR, hazard ratio; RCA, right coronary artery.

**Supplemental Table 4**. Unadjusted and adjusted Cox proportional hazard ratios in the external testing population for risk of major adverse cardiovascular events (MACE) with deep learning (DL) coronary artery calcium (CAC) scores.

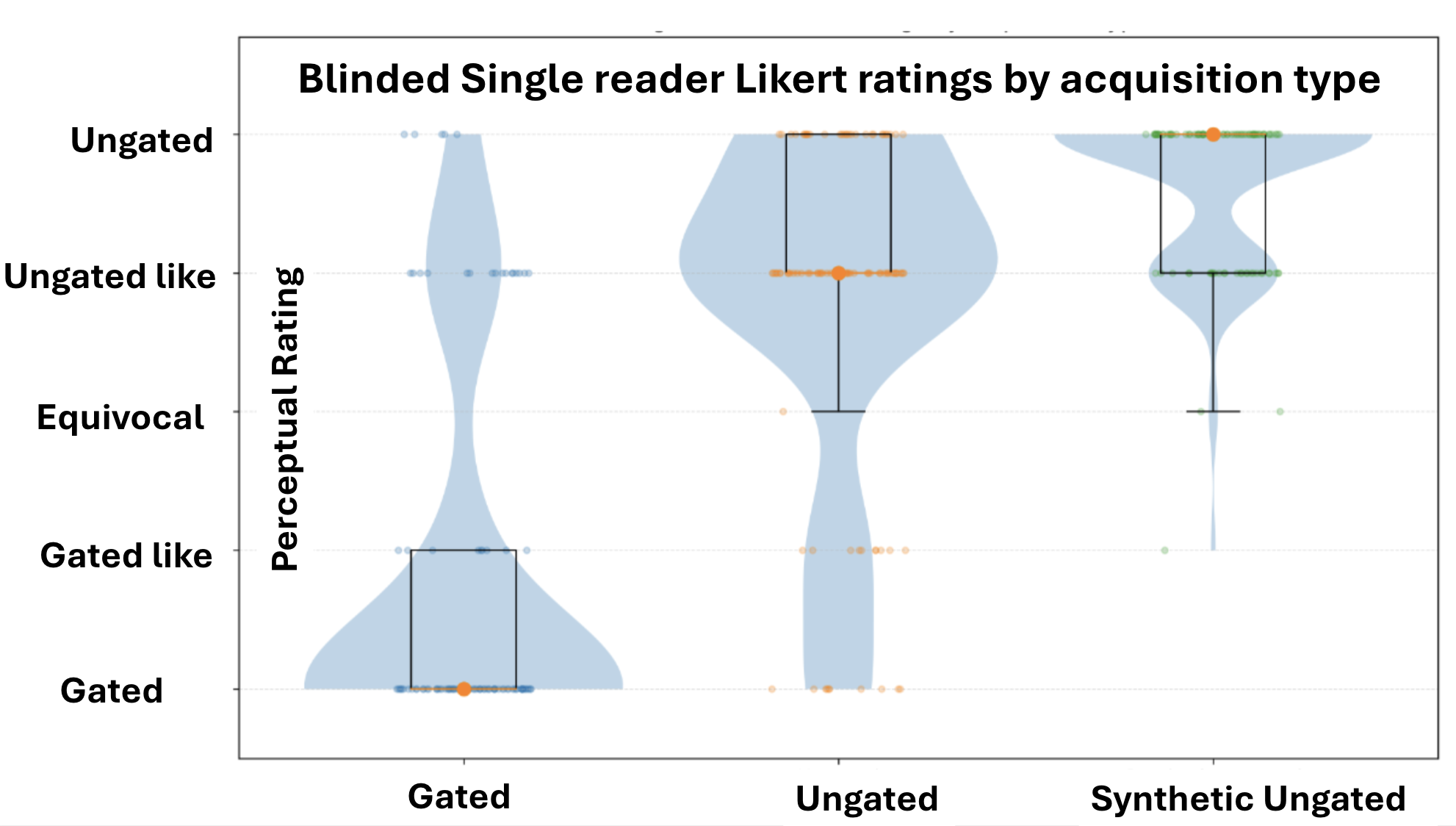

**Supplemental Figure 1.** Blinded expert-reader perceptual rating according to CT acquisition type (N = 300). Individual observations are shown as jittered points, with point colors used only to distinguish the three acquisition types (gated, ungated, and synthetic ungated CT). Violin width represents the relative density of ratings at each Likert category, such that wider regions indicate a greater concentration of observations at that rating. Superimposed boxplots summarize the distributions: the box represents the interquartile range, the central line represents the median, and the whiskers indicate the distributional spread.

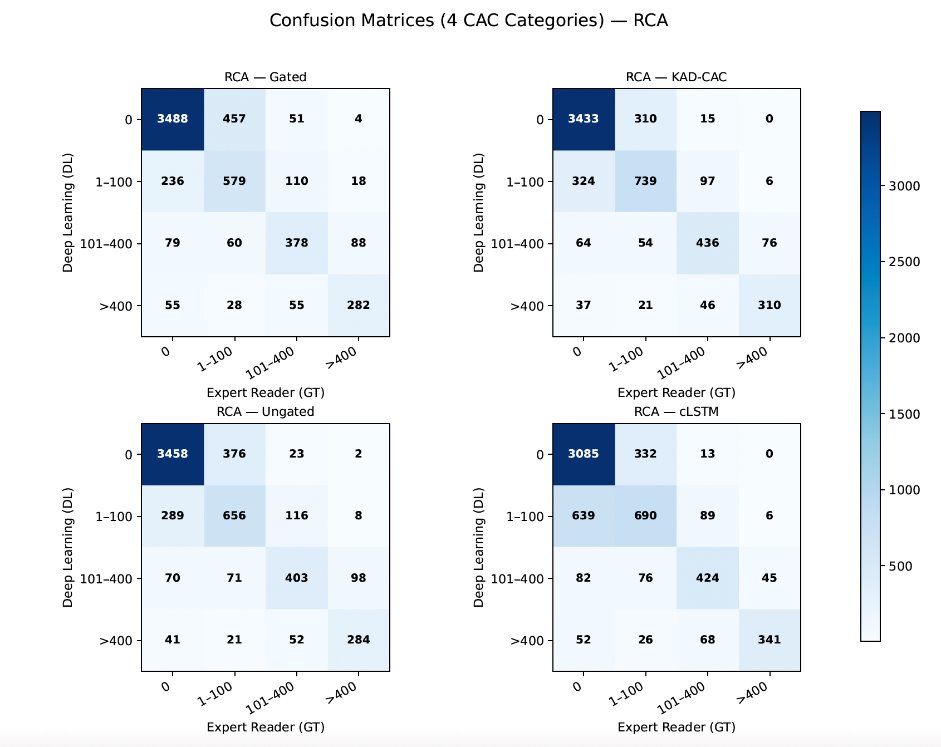

**Supplemental Figure 2:** Per-vessel confusion matrix for right coronary artery (RCA) coronary artery calcium scores.

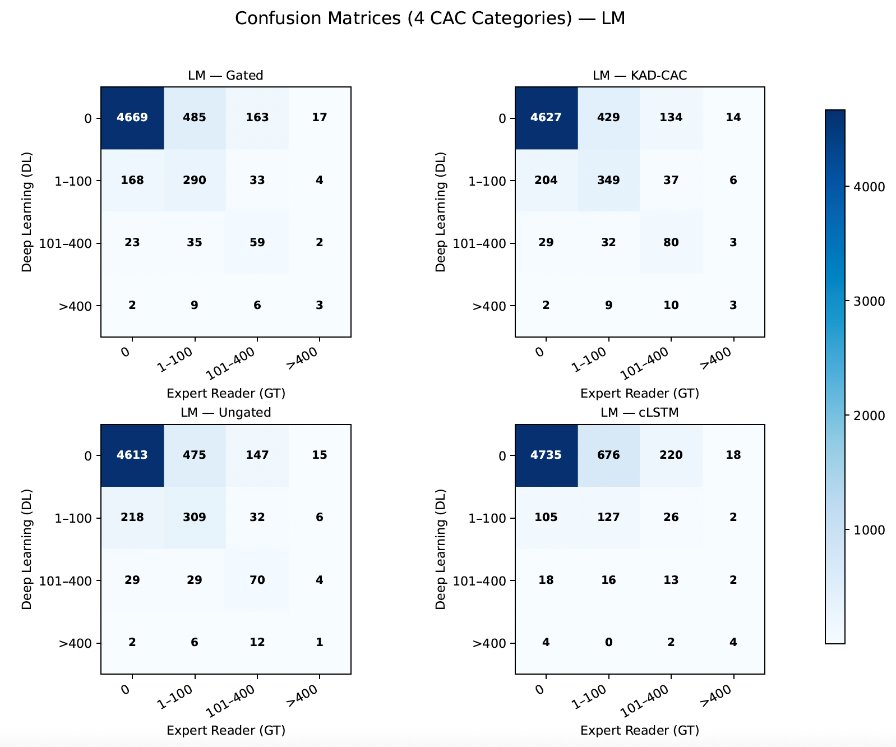

**Supplemental Figure 3:** Per-vessel confusion matrix for left main (LM) coronary artery calcium scores.

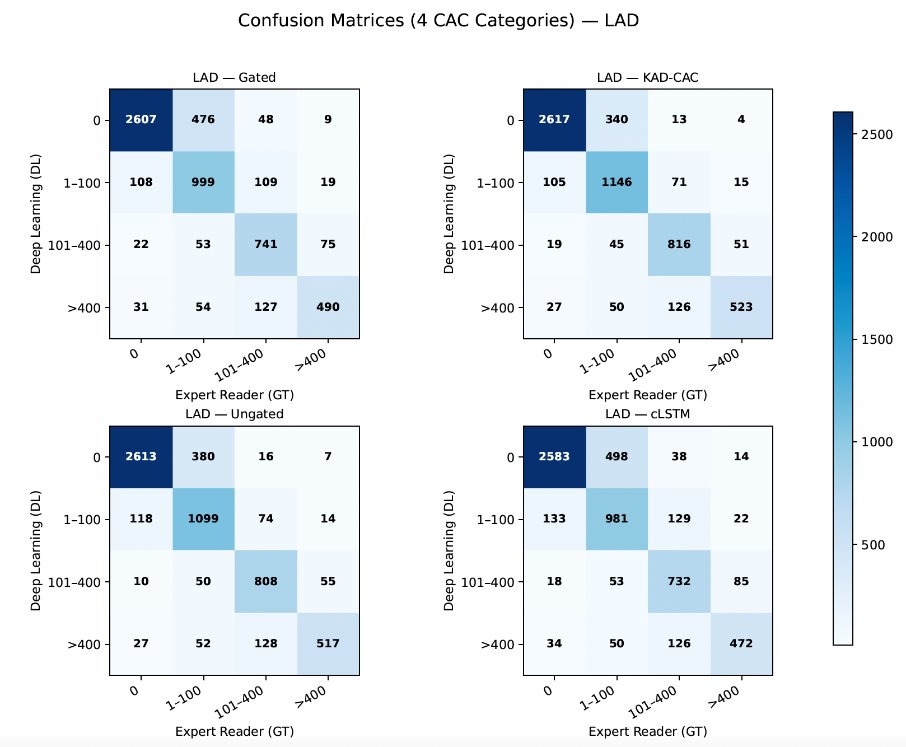

**Supplemental Figure 4:** Per-vessel confusion matrix for left anterior descending (LAD) coronary artery calcium scores.

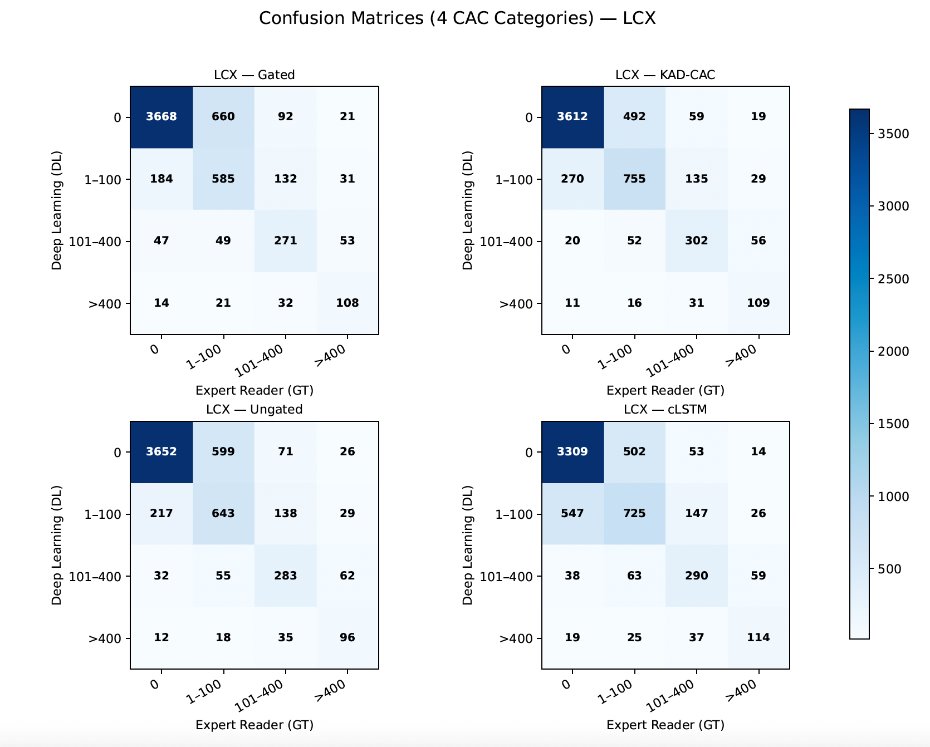

**Supplemental Figure 5:** Per-vessel confusion matrix for left circumflex (LCx) coronary artery calcium scores.

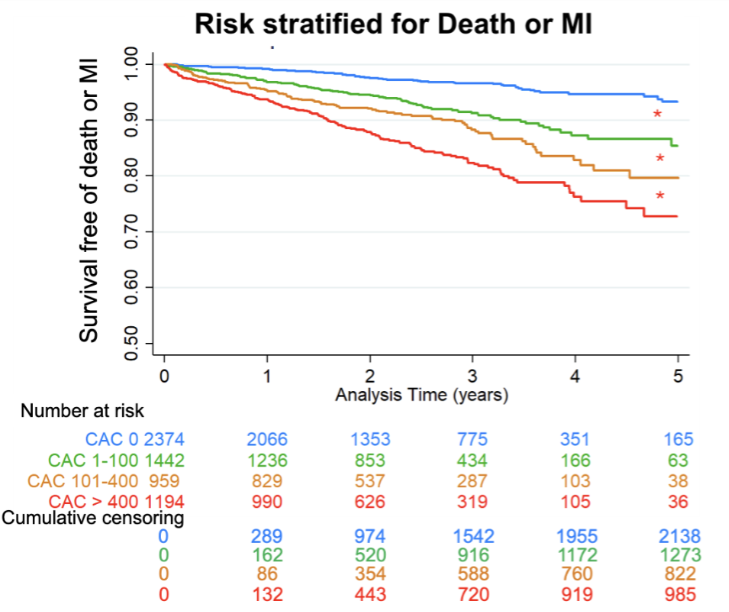

**Supplemental Figure 6.** Incidence of death or myocardial infarction (MI). Stratified by coronary artery calcification (CAC) as categorized by the KAD-CAC. * p<0.001

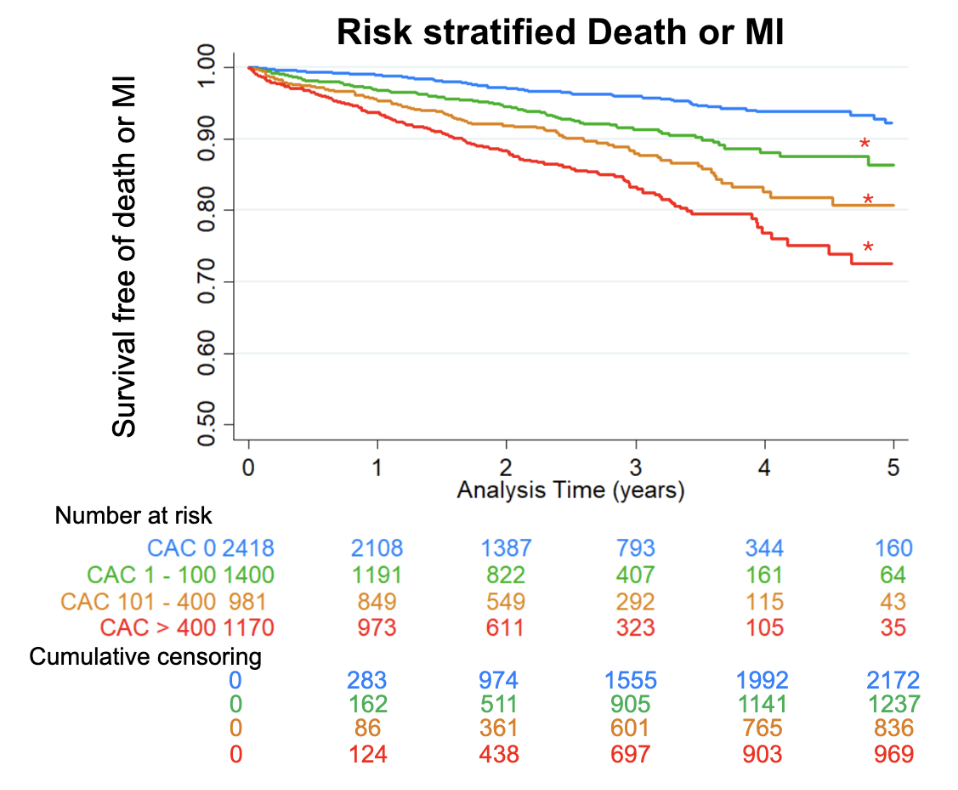

**Supplemental Figure 7.** Incidence of death or myocardial infarction (MI). Stratified by coronary artery calcification (CAC) as categorized by the expert reader scores. * p<0.001

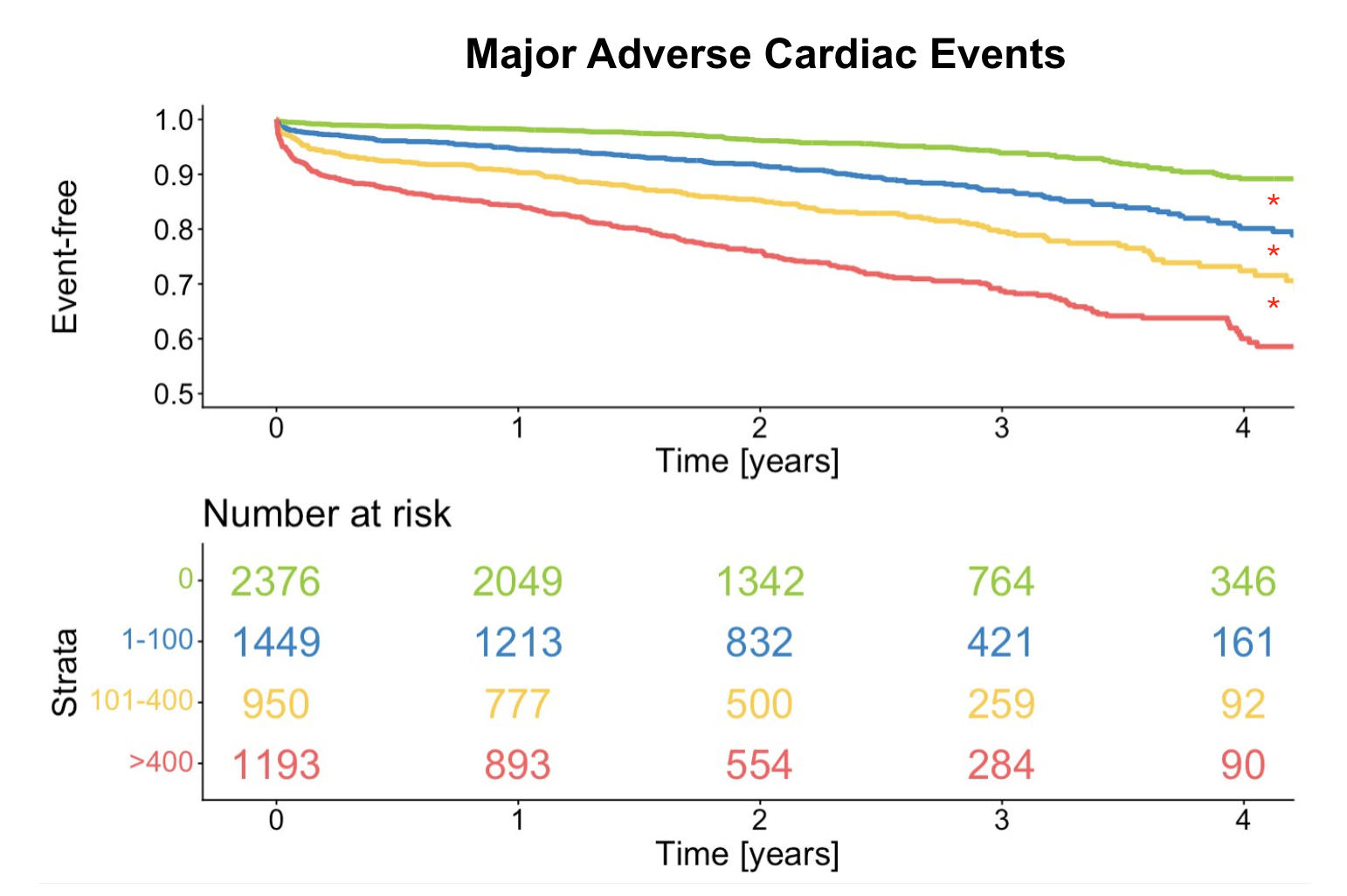

**Supplemental Figure 8.** Kaplan-Meier curves for the occurrence of MACE in patients with different severity of CAC assessed with DL on attenuation correction CT using KAD- CAC model. * p<0.01

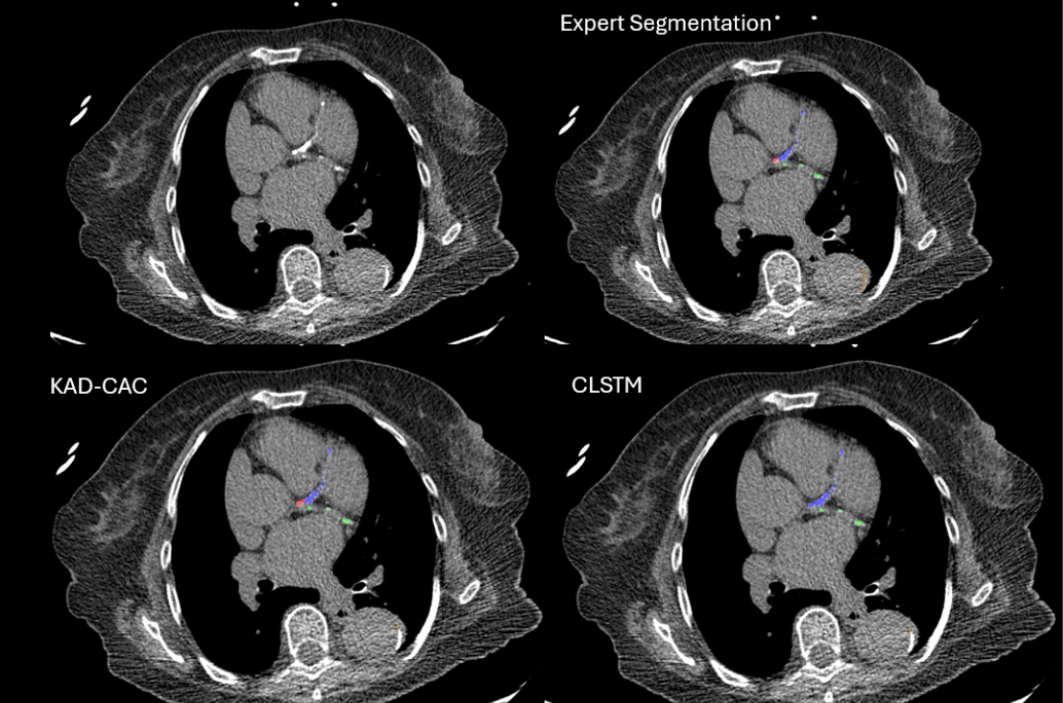

**Supplemental Figure 9.** Improvement in the vascular region CAC scoring in CTACs. Computed tomography attenuation correction (CTAC) from a female patient in early 80s. The expert left main (LM; colored red) coronary artery calcium (CAC) score was 41, and the KAD-CAC LM score was 49. The cLSTM model assigned the LM lesion to the LAD (blue) and LCx (green) territories, with LM score of 0.

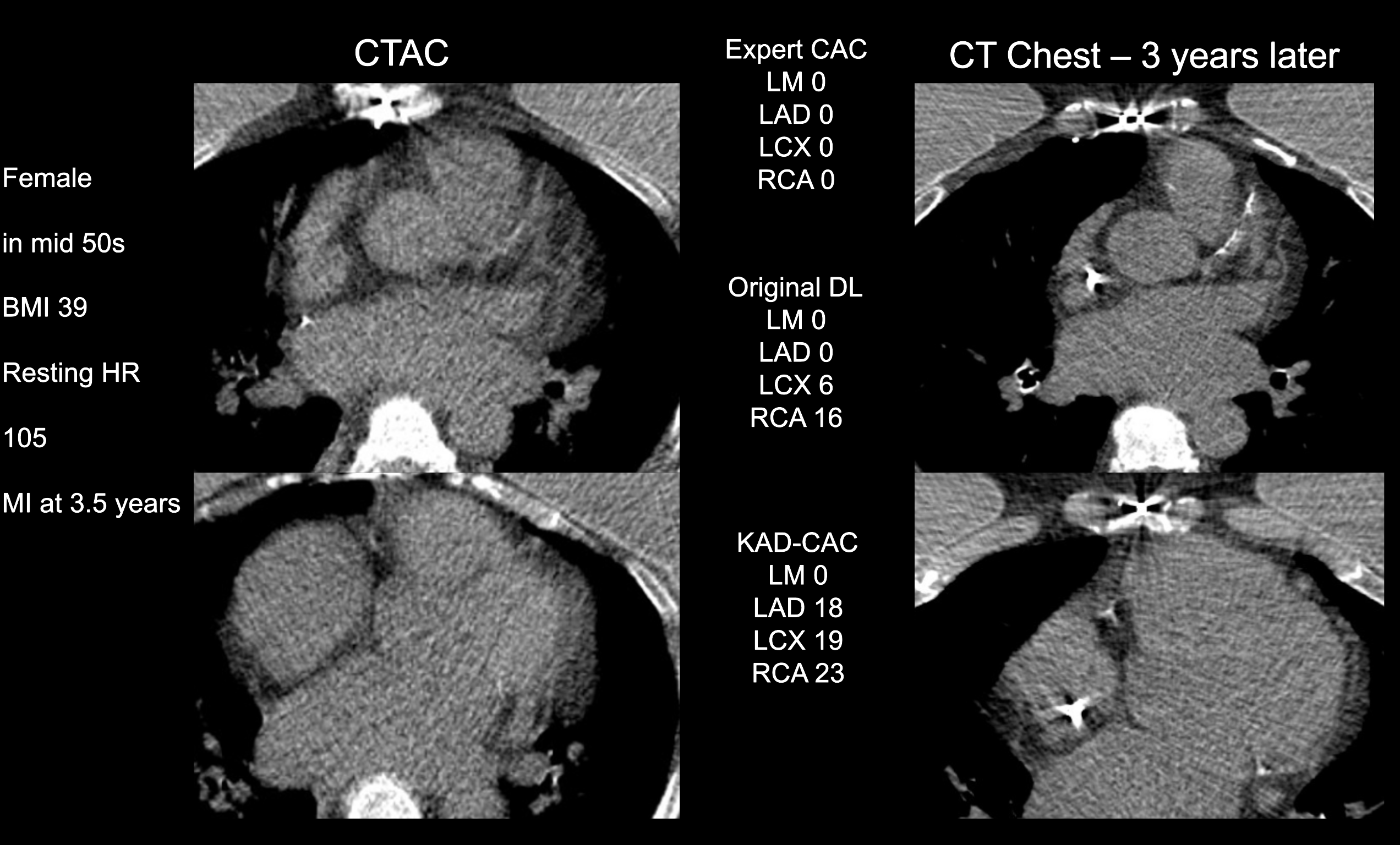

**Supplemental Figure 10:** Computed tomography attenuation correction (CTAC) imaging from a female patient in mid 50s. The expert CAC score was 0, likely related to poor image quality related to elevated body mass index (BMI) and heart rate. The KAD-CAC model identified three-vessel calcification. CT chest imaging performed three years later demonstrated three-vessel disease, with the patient experiencing MI at 3.5 years.

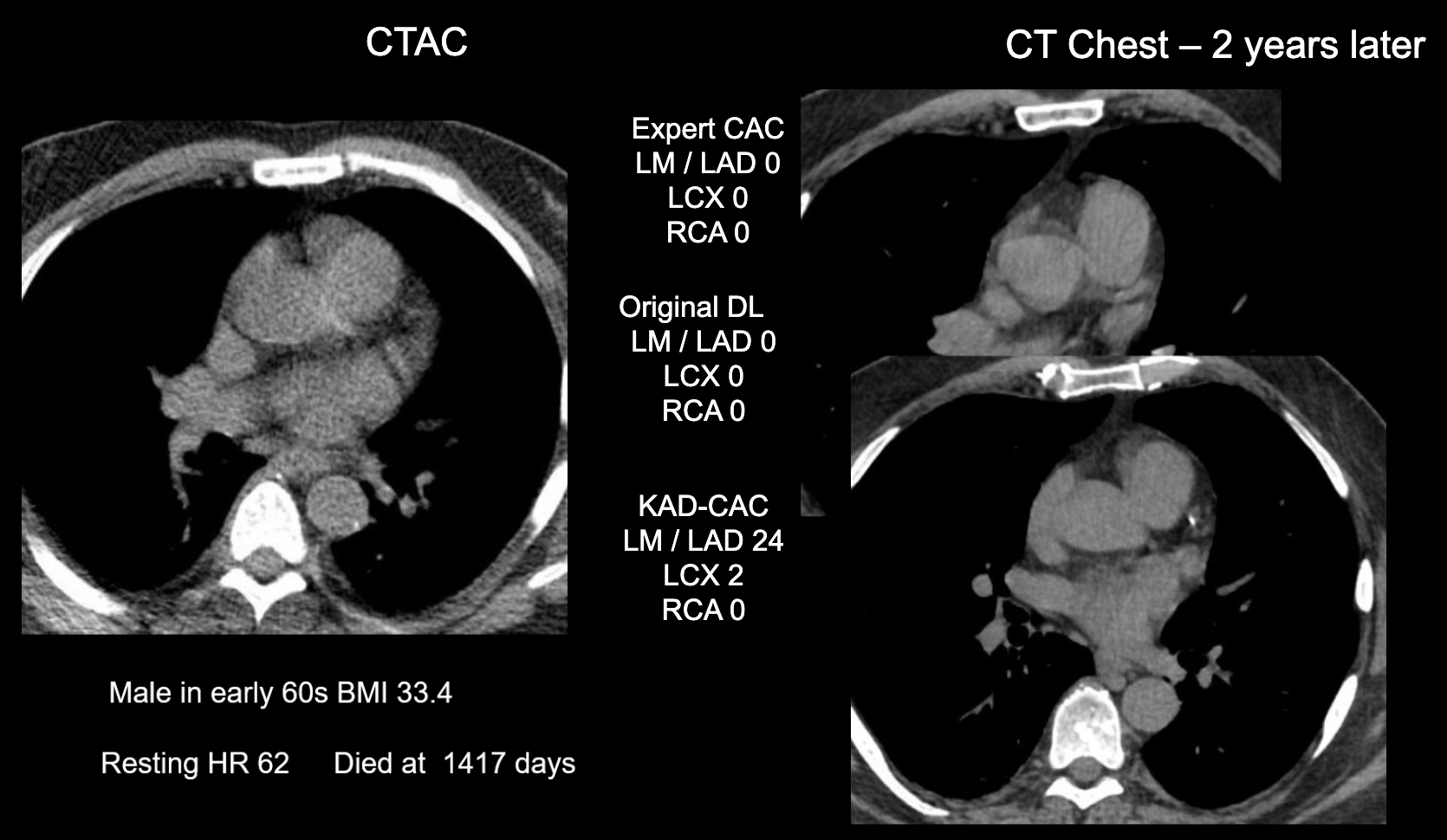

**Supplemental Figure 11:** Computed tomography attenuation correction (CTAC) imaging from a male in early 60s. The expert CAC score was 0. The KAD-CAC model identified LAD and LM disease that was more readily identified on a dedicated chest CT 2 years later.
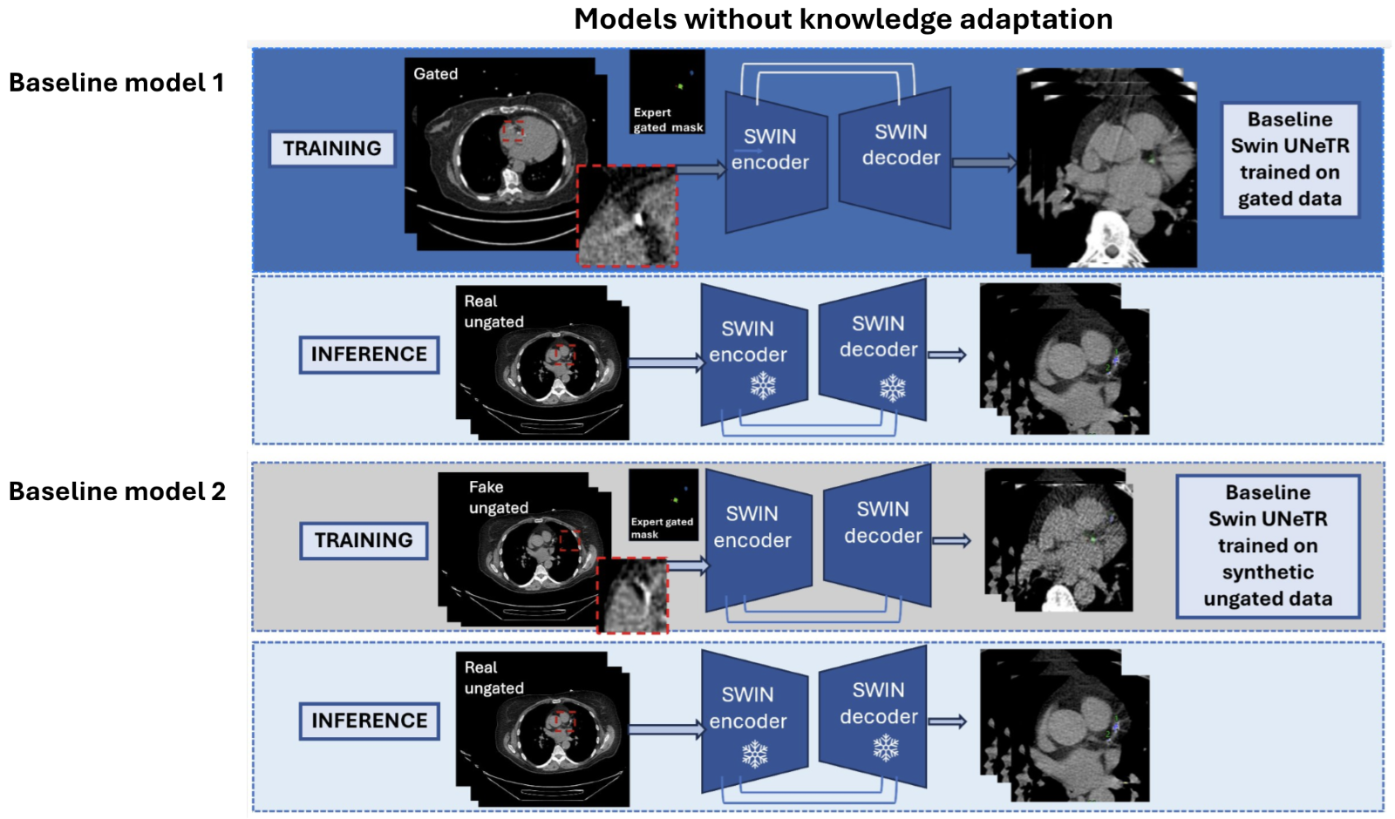
**Supplemental Figure 12.** Baseline transformer models for comparing against KAD-CAC model with cross domain knowledge transfer.

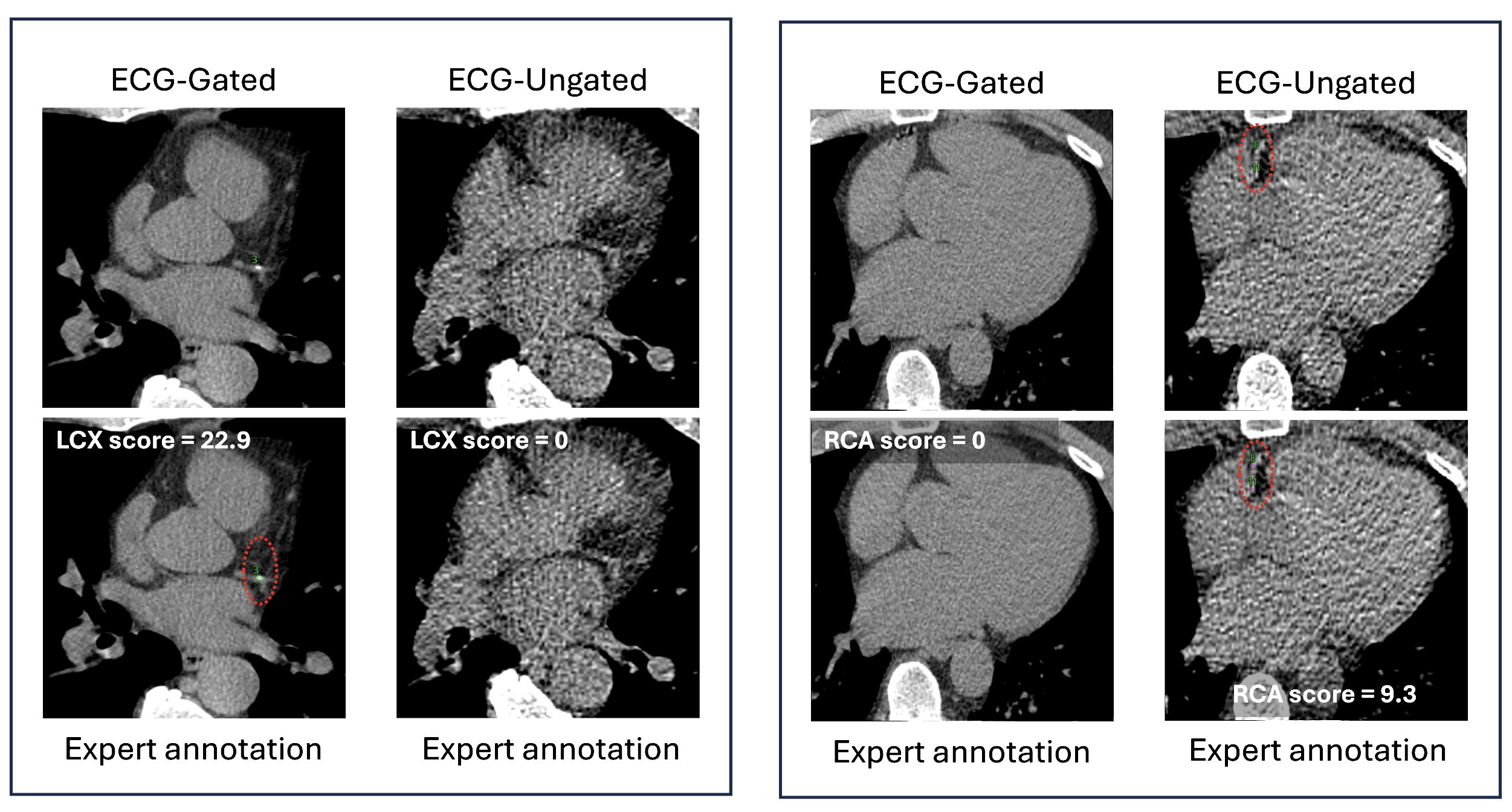

**Supplemental Figure 13.** Uncertainty in scoring for expert reader in paired gated CT and ungated CTAC due to respiratory motion, differences in breath-hold status, and partial volume effects, which can lead to loss or misregistration of coronary calcifications
